# High Dose Convalescent Plasma in COVID-19: Results from the Randomized Trial CAPSID

**DOI:** 10.1101/2021.05.10.21256192

**Authors:** Sixten Körper, Manfred Weiss, Daniel Zickler, Thomas Wiesmann, Kai Zacharowski, Victor M.Corman, Beate Grüner, Lucas Ernst, Peter Spieth, Philipp M. Lepper, Martin Bentz, Sebastian Zinn, Gregor Paul, Johannes Kalbhenn, Matthias Dollinger, Peter Rosenberger, Thomas Kirschning, Thomas Thiele, Thomas Appl, Benjamin Mayer, Michael Schmidt, Christian Drosten, Hinnerk Wulf, Jan Matthias Kruse, Bettina Jungwirth, Erhard Seifried, Hubert Schrezenmeier, for the CAPSID Clinical Trial Group

**Author notes:** Correspondence: Univ-Prof.Dr.med.Hubert Schrezenmeier, Institute for Clinical Transfusion Medicine and Immunogenetics Ulm, German Red Cross Blood Transfusion Service Baden-Württemberg-Hessen and University Hospital Ulm and Institute of Transfusion Medicine, University of Ulm. all members of the CAPSID Clinical Trial Group are listed in the Appendix. **Funding**: Bundesministerium für Gesundheit (German Federal Ministry of Health): ZMVI1-2520COR802. **Contributions**: HS and SK: Wrote study protocol, coordinated the study, analyzed and interpreted data, wrote the manuscript. HS, SK and ES applied for funding. HS and ES: Lead Investigators MW, DZ, TW, KZ, BG, LE, PS, PML, MB, SZ, GP, JK, MD, PR, TK, TT, HW, JMK, BJ: patient care, data collection VMC, CD: analysis of SARS-CoV-2 antibodies MS: SARS-CoV-2 PCR TA: project management BM: performed statistical analysis MS: SARS-CoV-2-PCR All authors have approved the manuscript.

## Abstract

**Rationale:** COVID-19 convalescent plasma (CCP) has been considered a treatment option in COVID-19.

**Objectives:** To assess the efficacy of neutralizing antibody containing high-dose CCP in hospitalized adults with COVID-19 requiring respiratory support or intensive care treatment.

**Methods:** Patients (n=105) were randomized 1:1 to either receive standard treatment and 3 units of CCP or standard treatment alone. Control group patients with progress on day 14 could cross over to the CCP group. Primary outcome was a dichotomous composite outcome of survival and no longer fulfilling criteria for severe COVID-19 on day 21. The trial is registered: clinicaltrials.gov #NCT04433910.

**Measurements and main results:** The primary outcome occurred in 43.4% of patients in the CCP and 32.7% in the control group (p=0.32). The median time to clinical improvement was 26 days (IQR 15-not reached (n.r.)) in the CCP group and 66 days (IQR 13-n.r.) in the control group (p=0.27). Median time to discharge from hospital was 31 days (IQR 16-n.r.) in the CCP and 51 days (IQR 20–n.r.) in the control group (p=0.24). In the subgroup that received a higher cumulative amount of neutralizing antibodies the primary outcome occurred in 56.0% (versus 32.1%), with a shorter interval to clinical improvement, shorter time to hospital discharge and better survival compared to the control group.

**Conclusion:** CCP added to standard treatment did not result in a significant difference in the primary and secondary outcomes. A pre-defined subgroup analysis showed a significant benefit for CCP among those who received a larger amount of neutralizing antibodies.

## Introduction

COVID-19 convalescent plasma (CCP) from patients recovered from a SARS-CoV-2 infection has become one of the treatment options for severe COVID-19. It has been broadly used in an Expanded Access Programm (1) in the US and preliminary reports on signals of efficacy and safety led to an Emergency Use Authorization in the US in August 2020 (2;3). A large number of clinical trials on CCP have been initiated since the start of the pandemic (4-16)(17). Efficacy has been mixed. A recent systematic review and meta-analysis concluded that CCP compared with placebo or standard of care was not significantly associated with a decrease in all-cause mortality or with any other benefit for other clinical outcomes (14). However, several limitations were noted: risk of bias, insufficient reporting of clinical outcomes other than all-cause mortality, and limited data to perform subgroup analyses. The certainty of evidence was considered low to moderate for all-cause mortality and low for other outcomes (14). The volume of CCP transfused was low in some of the trials (4;5;7;8;10;12) and the content of antibodies in CCP units was poorly characterized or only measured post-hoc in some of the trials. Therefore, it is important to gather data from further controlled clinical trials using well defined CCP.

Here we present the results of a randomized, prospective, open label, multicenter clinical trial of CCP compared with standard of care in hospitalized patients requiring supplemental oxygen or ventilation support or intensive care treatment (“CAPSID Trial”). It includes some unique features: Beside survival a series of other outcomes based on detailed definitions are reported. Patients in the CCP group received three units of plasma over a period of 5 days, i.e. a scheduled volume of about 850 ml CCP which is substantially higher than the CCP volume administered in other trials. Neutralizing SARS-CoV-2 antibodies were analyzed in detail both in the CCP products and in the recipients. Patients with progressive COVID-19 in the control group on day 14 could be switched to receive CCP.

## Methods

### Design

This is a multicenter, open-label randomized clinical trial to evaluate the efficacy and safety of CCP added to standard therapy (CCP group) vs. standard therapy alone (control group) in hospitalized patients with COVID-19 (**Figure 1**). Patients in the control group with progressive COVID-19 on day 14 were eligible to switch to treatment with CCP (crossover group). The trial was approved by the Federal Authority Paul-Ehrlich-Institute and by the Ethical Committee of the University of Ulm and the ethical committees of the participating hospitals. The trial is registered: EudraCT number 2020-001310-38 and NCT04433910. Written informed consent was obtained from all study participants or their legal representatives.

**Figure 1:**
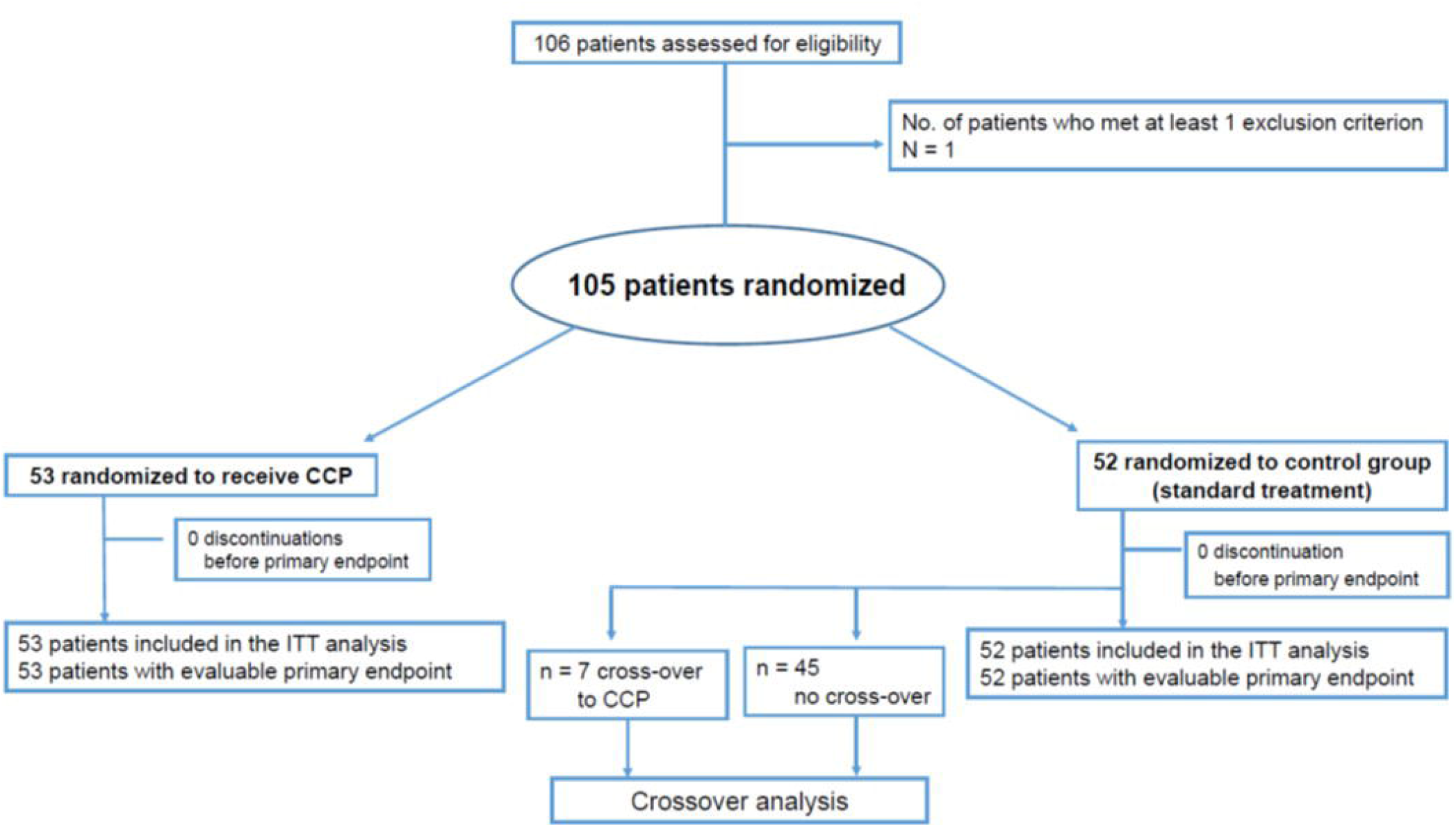
CONSORT Diagram.

### Patients

A total of 106 patients were recruited from 13 hospitals in Germany in the period from August 30, 2020 to December 24, 2020. Follow-up was completed on February 23, 2021. Inclusion criteria were: (1) SARS-CoV-2 infection confirmed by PCR (bronchoalveolar lavage, sputum, nasal and/or pharyngeal swap); (2) age ≥ 18 years and ≤ 75 years and (3) severe disease defined by at least one of the following: a) respiratory rate ≥ 30 breaths / minute under ambient air; b) requirement of any type of ventilation support (defined as supplemental oxygen or non-invasive ventilation or invasive ventilation or ECMO); c) needs treatment on ICU; (4) written informed consent by patient or representative. The full list of exclusion criteria is provided in the **Appendix**.

### Randomization

Patients (n=105) were randomized using a web-based system with a stratified 1:1 allocation ratio between each stratum (**Figure 1**). Patients were stratified prior to permutated block randomization by presence or absence of ventilation support, ECMO or ICU treatment.

### Crossover from Control Group to CCP treatment

Clinical condition in all patients was evaluated on day 14. In case of progression on day 14 compared to baseline, patients in the control group could receive CCP. A patient switching to CCP on day 14 was considered as failure of the primary outcome.

### SARS-CoV-2-Antibody Assays

A plaque reduction neutralization test (PRNT) and IgG and IgA-ELISA for SARS-CoV-2 were performed as previously described (18-20)

### Convalescent Plasma Transfusions and Standard treatment

The detailed characterization of CCP products has been reported elsewhere (21). The allocation of CCP to a recipient was based on the following criteria – provided availability: ABO-identical units, all three CCP units for a patient from one donor. If availability of CCP did not allow transfusing ABO-identical plasma, also minor compatible units were used. When all criteria were met the PRNT-50 titer was taken into account.

The administration of CCP should commence within 1 day after randomization. One transfusion unit each of CCP was given on day 1, 3 and 5. Since the total amount of neutralizing antibodies depends on both the volume and the antibody titer of CCP we used “neutralizing units” to take into account both variables. One neutralizing unit was arbitrarily defined as one ml of CCP with a PRNT50 titer of 1:20. The neutralizing units of a CCP transfusion unit were then calculated by dividing the titer by 20 and multiplying by volume (ml).

Patients in the crossover group also received one unit of CCP on three days.

Patients in both groups received other anti-viral treatment and/or supportive treatment according to institutional standard procedures.

### Outcome measures

The primary outcome of the CAPSID trial (to be interpreted as “treatment success”) was assessed on day 21 after randomization and is a dichotomous composite outcome of survival and no longer requiring ventilation support or ICU treatment and no tachypnea (i.e., respiratory rate <30 breaths/minute) on day 21. Key secondary outcomes were the time to clinical improvement and the frequency and severity of adverse events (AE). Clinical improvement was defined as an increase by at least two points on an ordinal severity scale (22). Patients without documented improvement were censored at last follow up. Further secondary outcomes were mortality, duration of ventilation support; time to discharge from ICU; time to hospital discharge; time until negative SARS-CoV-2 PCR from a nasopharyngeal swab

### Statistical Analysis

All patients were considered for the intention-to-treat (ITT) analysis independent from how they actually conducted the trial until assessment of the primary outcome (day 21).

Nominal and ordinal variables were analyzed by means of absolute frequencies and percentages. Missing values were considered as a separate category. Continuous variables are described by presenting the median and interquartile range (IQR) for the total number of patients who contributed values. For PRNT50, the geometric mean, variance and 95% confidence intervals (CI) are presented. The primary outcome was analyzed using Fisher’s exact test comparing the treatment success rates in both treatment groups. A two-sided p-value of <0.05 will be considered statistically significant.

Post-hoc analyses were added to compare outcome in patients with presence or absence of neutralizing antibodies at baseline and patients with or without invasive ventilation at baseline.

No imputation was necessary for primary outcome. Missing data for secondary outcomes and adverse events were not imputed. All statistical analyses were performed according to the statistical analysis plan using SAS® (version 9.4M6 or newer, www.sas.com).

More details of methods are provided in the **Appendix**.

## Results

### Study population

A total of 106 patients were enrolled to this clinical trial and 105 patients were randomized to either the CCP group (n=53) or the control group (n=52)(**Figure 1**) All 105 patients were included in the intention-to-treat analysis.

Baseline demographics and clinical characteristics are shown in **Table 1**. The majority of patients were male (73.3%). The median age was 60 years (IQR 53-66). A majority of patients (89.5%) had a coexisting condition at entry into the trial (56.2% hypertension, 31.4% diabetes, 21.9% cardiovascular disease). The median body mass index was 29.4 kg/m^2^. The percentage of patients receiving supplemental oxygen or non-invasive ventilation (score 4 and 5 on the ordinal severity scale) or invasive ventilation (score 6 and 7) was 59.1% and 34.3%. The time from symptom onset of the SARS-CoV-2 infection to randomization was 7 days (IQR 4-10). SARS-CoV-2 PCR from nasopharyngeal swabs was still positive in 94.3% of patients at baseline. Neutralizing SARS-CoV-2 antibodies were present in 78.9% of patients with available information at baseline (median titer among those with detectable antibodies 1:160 (IQR 1:80 – 1:640).

**Table 1:** Baseline Demographics and Clinical Characteristics.

|  | CCP group<br>(n=53) | Control Group<br>(n=52) | p-value |
| --- | --- | --- | --- |
| **Demographic and clinical characteristics** |  |  |  |
| Median age, years (IQR) | 59 (53-65) | 62 (55-66) | 0.24 |
| Gender, no (%) |  |  | 0.19 |
| Female | 11 (20.8) | 17 (32.7) |  |
| Male | 42 (79.3) | 35 (67.3) |  |
| Body Mass Index, kg/m<sup>2</sup> (IQR) | 29.4 (27.6-33.4) | 29.1 (25.6-31.5) | 0.07 |
| Coexisting Diseases, n (%) |  |  |  |
| no other disease | 7 (13.2) | 4 ( 7.7) |  |
| BMI >30 kg/m<sup>2</sup> | 28(52.8) | 29 (55.6) |  |
| Hypertension | 31 (58.5) | 28 (53.9) |  |
| Cardiovascular disease | 12 (22.6) | 11 (21.2) |  |
| Diabetes | 18 (34.0) | 15 (28.9) |  |
| COPD, Asthma, other<br>pulmonary disease | 8 (15.1) | 9 (17.3) |  |
| Thromboembolic disease | 2 ( 3.8) | 3 ( 5.8) |  |
| solid tumor | 2 ( 3.8) | 3 ( 5.8) |  |
| other | 32 (60.4) | 41 (78.8) |  |
| Point Scale at Study entry, n (%) |  |  | 0.68 |
| 3 | 4 ( 7.6) | 3 ( 5.8) |  |
| 4 | 5 ( 9.4) | 8 (15.4) |  |
| 5 | 28 (52.8) | 21 (40.4) |  |
| 6 | 3 ( 5.7) | 3 ( 5.8) |  |
| 7 | 13 (24.5) | 17 (32.7) |  |
| Respiratory Rate,<br>breaths/min (IQR) | 24 (20-30) | 23 (18-30) | 0.82 |
| Median time from symptom onset of<br>the SARS-CoV-2 infection to<br>randomization, days (IQR) | 7 (2-9) | 7 (5-10.5) | 0.07 |
| Median time from hospitalization to<br>randomization, days (IQR)<sup>§</sup> | 1 (1-3) | 2.5 (1-5) | 0.02 |
| Prior /Concomitant medication, n (%) |  |  |  |
| Antiviral drug | 23 (43.4) | 24 (46.2) |  |
| Corticosteroids | 45 (84.9) | 49 (94.2) |  |
| Antibiotic drug | 26 (47.2) | 25 (48.1) |  |
| Vasopressors | 24 (45.3) | 30 (57.7) |  |
| Anticoagulation | 42 (79.3) | 40 (76.9) |  |
| Platelet aggregation inhibitor | 17 (32.1) | 15 (28.9) |  |
| RBC Transfusion | 14 (26.4) | 25 (48.1) |  |
| PLT Transfusion | 1 ( 1.9) | 4 ( 7.7) |  |
| FFP Transfusion | 7 (13.2) | 9 (17.3) |  |
| No concomitant medication | 2 ( 3.8) | 2 ( 3.9) |  |
| **SARS-CoV-2 status at baseline** |  |  |  |
| Result of SARS-CoV-2 PCR<br>nasopharyngeal swab, n (%) |  |  | 0.62 |
| positive | 49 (92.5) | 50 (96.2) |  |
| negative | 3 ( 5.7) | 1 ( 1.9) |  |
| missing | 1 ( 1.9) | 1 ( 1.9) |  |
| **Humoral immune response at baseline** |  |  |  |
| **SARS-CoV-2 IgA present, n (%)** |  |  | 0.58 |
| Yes | 37 (69.8) | 38 (73.1) |  |
| No | 6 (11.3) | 7 (13.5) |  |
| Missing | 10 (18.9) | 7 (13.5) |  |
| **SARS-CoV-2 IgG present, n (%)** |  |  | 0.50 |
| Yes | 25 (47.2) | 30 (57.7) |  |
| No | 19 (35.9) | 15 (28.9) |  |
| Missing | 9 (17.0) | 7 (13.5) |  |
| **PRNT50 Titer, Categorical, n (%)** |  |  | 1.00 |
| Negative | 10 (18.9) | 10 (19.2) |  |
| 1: 20 | 2 ( 3.8) | 2 ( 3.9) |  |
| 1: 40 | 5 ( 9.4) | 6 (11.5) |  |
| 1: 80 | 5 ( 9.4) | 7 (13.5) |  |
| 1:160 | 6 (11.3) | 5 ( 9.6) |  |
| 1:320 | 7 (13.2) | 7 (13.5) |  |
| 1:640 | 5 ( 9.4) | 4 ( 7.7) |  |
| >1:640 | 7 (13.2) | 7 (13.5) |  |
| Missing | 6 (11.3) | 4 ( 7.7) |  |
| **Neutralizing antibodies (based on PRNT50 titer ≥1:20) present** |  |  | 1.00 |
| no | 10 (18.9) | 10 (19.2) |  |
| yes | 37 (69.8) | 38 (73.8) |  |
| Missing | 6 (11.3) | 4 ( 7.7) |  |
| Median (IQR)* | 1:320 (1:80-1:640) | 1:160 (1:80-1:640) | 0.85 |
| Mean (SD)* | 1:432 (1:457) | 1:405 (1:458) |  |
| **Laboratory values at baseline** |  |  |  |
| **Inflammation Markers** |  |  |  |
| Ferritin [µg/L], median (IQR) | 1040 (649-1552) | 1185 (711-2200) | 0.01 |
| CRP [mg/L], median (IQR) | 151 (79-230) | 130 (66-206) | 0.74 |
| IL-6 [pg/ml], median (IQR) | 60 (16-98) | 44 (18-142) | 0.40 |
| LDH [U/L], median (IQR) | 481 (399-551) | 486 (362-662) | 0.74 |
| **Renal and hepatic markers** |  |  |  |
| Serum Creatinine [mg/dl], median (IQR) | 0.96 (0.74-1.23) | 1.09 (0.80-1.65) | 0.07 |
| Bilirubin [mg/dl], median (IQR) | 0.47 (0.32-0.62) | 0.51 (0.30-0.81) | 0.15 |
| **Blood Count** |  |  |  |
| White blood cells [x10<sup>9</sup>/L], median (IQR) | 8.23 (6.88-11.3) | 9.92 (7.03-14.03) | 0.11 |
| Lymphocytes [x10<sup>9</sup>/L], median (IQR) | 0.68 (0.40-0.80) | 0.54 (0.30-0.83) | 0.82 |
| Hemoglobin [g/dL], median (IQR) | 13.0 (11.7-14.2) | 12.0 (10.4-14.0) | 0.88 |
| Platelets [x10<sup>9</sup>/L], median (IQR) | 231 (195-320) | 237 (163-301) | 0.83 |

\*this descriptive analysis included only patients with detectable antibodies (PRNT50 ≥ 1:20).
RBC, Red blood cell concentrate; PLT platelet concentrate; FFP – fresh frozen plasma (non-immune plasma, not for indication of passive immunotherapy against SARS-CoV-2).

Overall, the CCP group and the control group were similar in terms of demographic characteristics and disease severity as assessed by the distribution on the ordinal severity scale, the type of ventilation support and the laboratory results at baseline.

### Study Treatment

Fifty-two out of 53 (98.1%) patients randomized to CCP received all three planned CCP transfusions with a median total volume of 846 ml (IQR 824-855 ml). The median PRNT50 neutralization titer of CCP was 1:160 (IQR 1:80 to 1:320)(**Appendix**, Figure 1). The median transfused neutralizing units per patients were 6768 (IQR 3424-13520). The majority of patients (96.2%) received only ABO-identical transfusions, one patient each with type A and type B received AB plasma, and 94.3% received all three plasma units from one donor collected in either one or two plasmapheresis sessions (79.3% and 15.1%, resp). Crossover patients received a median total volume of 837 ml (IQR 738 ml – 872 ml) and all but one crossover patients received all three CCP transfusions.

### Primary Clinical Outcome

There was no significant difference in the dichotomous composite primary outcome at day 21 (no longer requiring ventilation support or ICU treatment and no tachypnea, i.e. respiratory rate < 30/minute): 43.4% of patients in the CCP group and 32.7% in the control group reached the primary outcome (p=0.32) (**Table 2a**). Among those who received a low or high cumulative amount of neutralizing units the primary outcome occurred in 32.1% and 56.0% (**Table 2b**). Among those with low and high inflammation markers at entry, the primary outcome occurred in 54.2% and 25.5% (**Table 2c**) (p=0.004, Fisher’
ss exact test; explorative analysis). Among the patients without or with neutralizing antibodies at baseline, the primary outcome occurred in 25.0% and 42.7% (p=0.20, Fisher’s exact test, explorative analysis)(**Table 2d**). Among those with or without invasive ventilation or ECMO at entry the primary outcome occurred in 13.9% and 50.7% (p<0.001, Fisher’s exact test, explorative analysis)(**Table 2e**). However, within the two subgroups the outcome did not differ between the CCP group and the control group.

**Table 2a:**
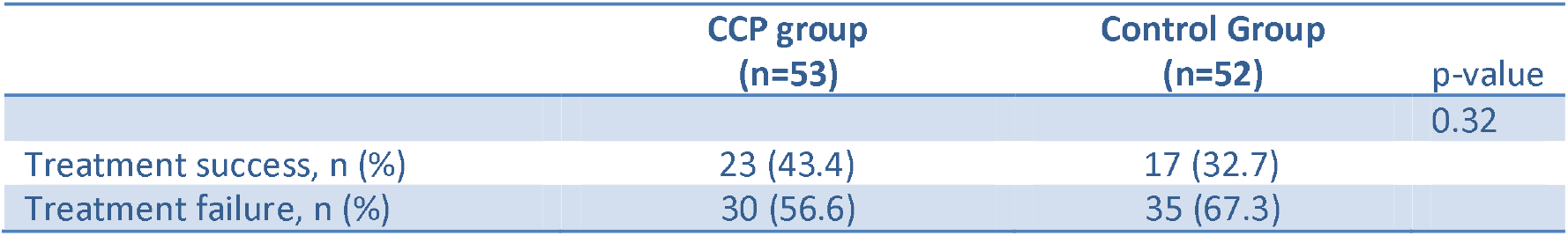
Primary Outcome at Day 21.

|  | CCP group<br>(n=53) | Control Group<br>(n=52) | p-value |
| --- | --- | --- | --- |
|  |  |  | 0.32 |
| Treatment success, n (%) | 23 (43.4) | 17 (32.7) |  |
| Treatment failure, n (%) | 30 (56.6) | 35 (67.3) |  |

**Table 2b:**
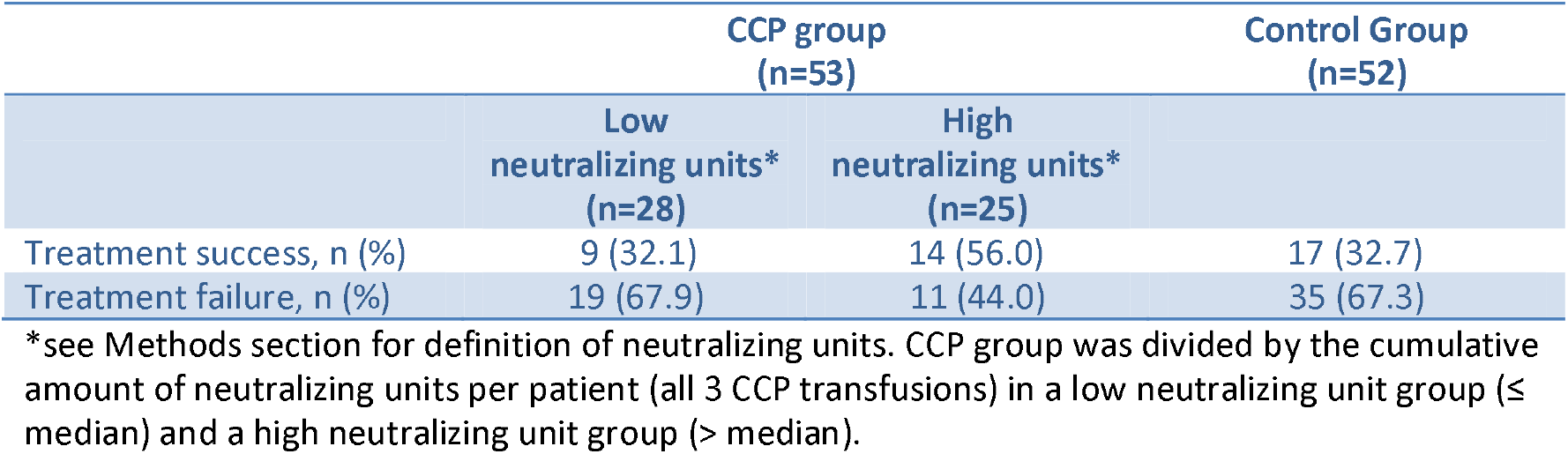
Primary Outcome at Day 21 by Transfused Neutralizing Units*.

**Table 2c:** Primary Outcome at Day 21 by Inflammation Markers*.

|  | High Inflammation Markers<br>(n=51)** |  | Low inflammation Markers<br>(n=48)** |  |
| --- | --- | --- | --- | --- |
|  | CCP Group<br>(n=25) | Control Group<br>(n=26) | CCP Group<br>(n=24) | Control Group<br>(n=24) |
| Treatment success, n (%) | 8 (32.0) | 5 (19.2) | 14 (58.3) | 12 (50.0) |
| Treatment failure, n (%) | 17 (68.0) | 21 (80.8) | 10 (41.7) | 12 (50.0) |

\*see Methods section for definition of high / low inflammation markers. The patient group was divided in a low inflammation marker group and a high neutralizing unit group. In each of these groups the primary outcome for patients in the CCP group and the control group was compared.
\*\* Six patients with either missing data on inflammation markers (n=1) or intermediate inflammation markers (n=5) are not included in this table.

**Table 2d:** Primary Outcome at Day 21 by Presence / Absence of SARS-CoV-2 Neutralizing Antibodies at Baseline *.

|  | anti-SARS-CoV-2 neutralizing<br>antibodies at baseline:<br>positive<br>(n=75)** |  | anti-SARS-CoV-2 neutralizing<br>antibodies at baseline:<br>negative<br>(n=20)** |  |
| --- | --- | --- | --- | --- |
|  | CCP Group<br>(n=37) | Control Group<br>(n=38) | CCP Group<br>(n=10) | Control Group<br>(n=10) |
| Treatment success, n (%) | 17 (46.0) | 15 (39.5) | 4 (40.0) | 1 (10.0) |
| Treatment failure, n (%) | 20 (54.0) | 23 (60.5) | 6 (60.0) | 9 (90.0) |

\* presence of anti-SARS-CoV-2 neutralizing antibodies PRNT50 ≥ 1:20 at baseline.
\*\* Ten patients with missing data on neutralizing antibodies (PRNT50) at baseline are not included in this table.

**Table 2e:** Primary Outcome at Day 21 by Ventilation Status at Baseline.

|  | Patients without<br>invasive ventilation / ECMO<br>(n=69) |  | Patients with<br>invasive ventilation / ECMO<br>(n=36) |  |
| --- | --- | --- | --- | --- |
|  | CCP Group<br>(n=37) | Control Group<br>(n=32) | CCP Group<br>(n=16) | Control Group<br>(n=20) |
| Treatment success, n (%) | 21 (56.8) | 14 (43.8) | 2 (12.5) | 3 (15.0) |
| Treatment failure, n (%) | 16 (43.2) | 18 (56.3) | 14 (87.5) | 17 (85.0) |

### Secondary Clinical Outcome

The median time to clinical improvement by ≥ 2 points on the ordinal severity scale was 26 days (IQR 15-n.r.) in the CCP group and 66 days (IQR 13-n.r.) in the control group (p= 0.27, log-rank test)(**Figure 2A**). The majority of patients (81.0%) experienced at least one AE. Neither the frequency of AEs nor the worst AE grade did significantly differ between the groups (p=0.62 and p=0.18, resp.)(**Appendix**, Table 1). The outcome of AEs was similar between the groups. The proportion of patients with serious AEs (SAEs) was 41.5% in the CCP group and 48.1% in the control group. Number of SAEs and reasons for classification as SAE are summarized in **Appendix**, Table 1.

**Figure 2:**
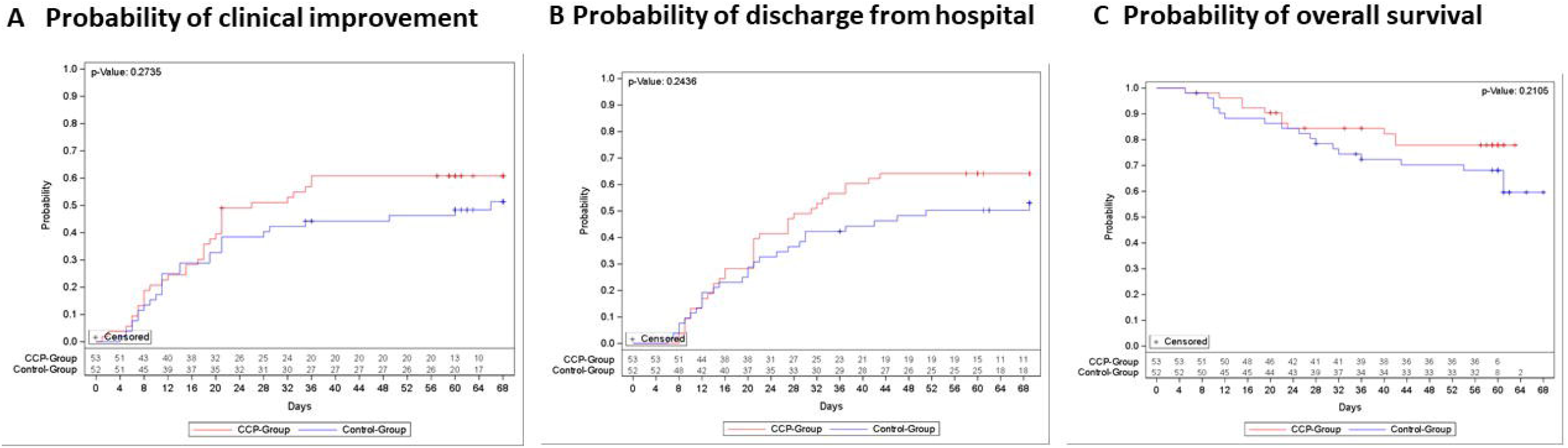
Occurrence of Secondary Outcomes. Kaplan-Meier cumulative estimates of probability of (A) the key secondary outcome time to clinical improvement compared in the CCP group (red line) or control group (blue line). Censored patients are indicated by +. p=0.27 (log-rank test). (B) discharge from hospital compared in the CCP group (red line) or control group (blue line). Censored patients are indicated by +. p= 0.24 (log-rank test). (C) overall survival compared in the CCP group (red line) or control group (blue line). Censored patients are indicated by +. p=0.21 (log-rank test).

The case fatality rate at day 21, day 35 and day 60 was 14.3%, 21.0% and 26.7%, resp., without a significant difference between the groups (**Appendix**, Table 2). The probability of overall survival at day 60 is 77.9% (95%-CI 63.6-87.1%) in the CCP group and 68.1% (95%-CI 53.3%-79.1%) in the control group (p=0.21, log-rank test) (**Figure 2C**).

The distribution of clinical outcomes according to the ordinal severity scale is shown in **Figure 4**. On day 35, 65.3% of patients with available follow-up information in the CCP group and 44.0% in the control group were discharged from hospital or did no longer require supplemental oxygen (p=0.04, Fisher’
ss exact test, exploratory analysis). The proportion of patients still requiring supplemental oxygen, non-invasive or invasive ventilation on day 35 was 18.4% in the CCP group and 28.0% in the control group. The median time to discharge from ICU was 29 days (IQR 9-n.r.) in the CCP group and 42 days (IQR 12-n.r.) in the control group (p=0.39, log-rank test)(**Figure 5A**). The median time to discharge from hospital was 31 days (IQR 16-n.r.) in the CCP group and 51 days (IQR 20-n.r.) in the control group (p=0.24, log-rank test)(**Figure 2B**).

The median time to first negative SARS-CoV-2 PCR from nasopharyngeal swab was 7 days (IQR 4-17 days) in the CCP group and 8 days (IQR 5-21 days) in the control group (p=0.38, log-rank test)(**Appendix**, Figure 2a).

The secondary time to event outcomes by transfused neutralizing units are summarized in **Appendix**, Table 3. Among those patients in the CCP group who received a high cumulative amount of neutralizing units when compared to those with a low amount the median time to clinical improvement was 20 days (IQR 11-n.r.) and 36 days (IQR 17-n.r.)(**Figure 3A; Appendix**, Table 3). The median time do discharge from hospital was 21 days (IQR 13-43) and 39 days (IQR 21-n.r.)(**Figure 3B; Appendix**, Table 3), the median time to discharge from ICU was 14 days (7-39) and 39 days (20-n.r.)(**Figure 5B; Appendix**, Table 3) and the median time to negative SARS-CoV-2 PCR was 5 days (IQR 3-15) and 14 days (IQR 5-19)(**Appendix**, Figure 2b and Table 3). Overall survival was higher among patients transfused with high amounts of neutralizing antibodies compared to the control group (p=0.02)(**Figure 3C**).

**Figure 3:**
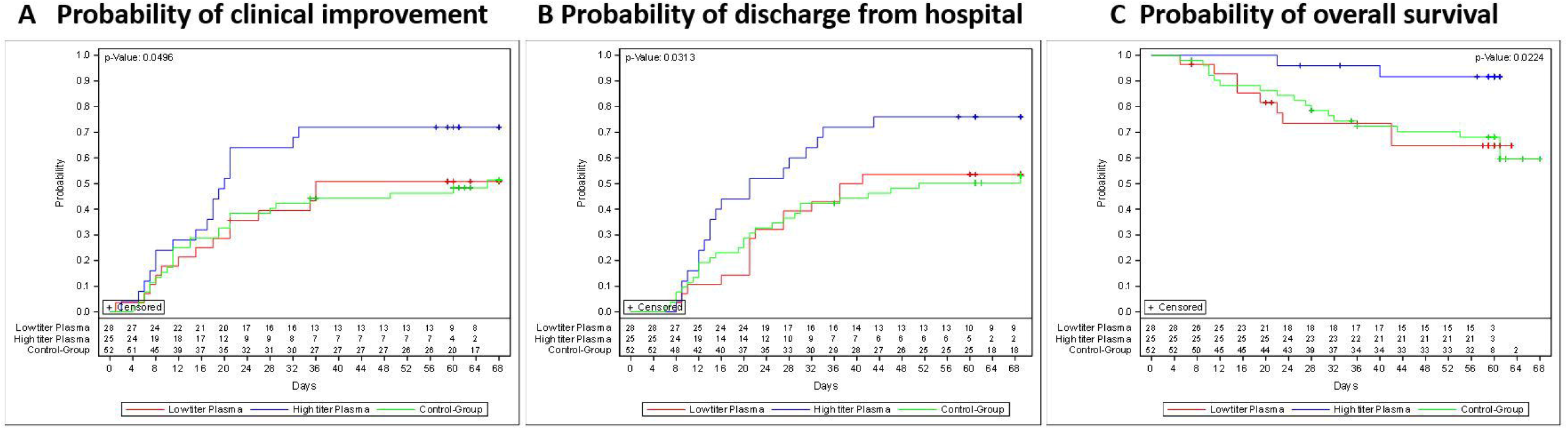
Occurrence of Secondary Outcomes by Cumulative Amount of Transfused Neutralizing Units. Kaplan-Meier cumulative estimates of probability of (A) the key secondary outcome time to clinical improvement compared in the CCP subgroup which received a low cumulative amount of neutralizing units (red), the CCP subgroup which received a high cumulative amount of neutralizing units (blue) and the control group (green line). Censored patients are indicated by +. p=0.05 (log-rank test; high amount vs. control group). (B) discharge from hospital compared in the CCP subgroup which received a low cumulative amount of neutralizing units (red), the CCP subgroup which received a high cumulative amount of neutralizing units (blue) and the control group (green line). Censored patients are indicated by +. p=0.03 (log-rank test, high amount vs. control group)). (C) overall survival compared in the CCP subgroup which received a low cumulative amount of neutralizing units (red), the CCP subgroup which received a high cumulative amount of neutralizing units (blue) and the control group (green line). Censored patients are indicated by +. p=0.02 (log-rank test, high amount vs. control group).

**Figure 4:**
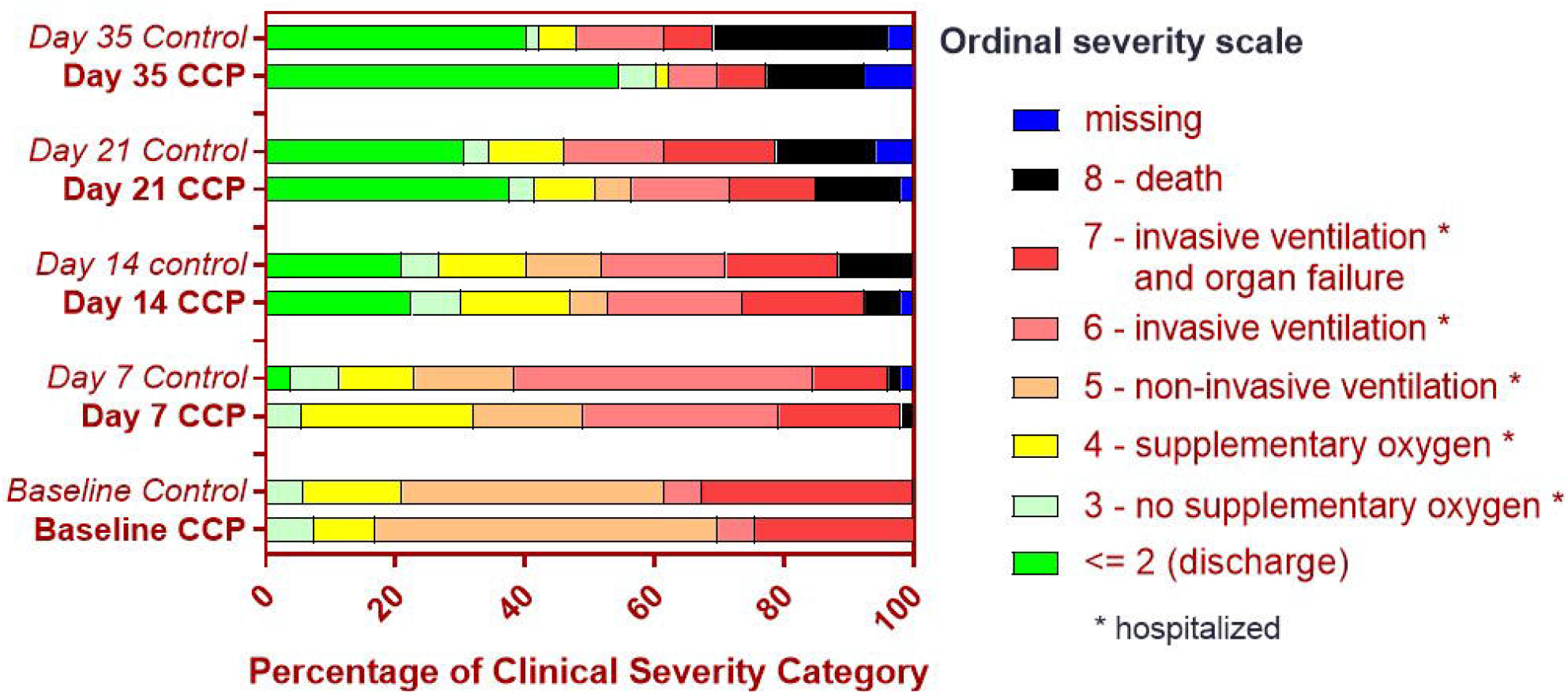
Clinical Outcomes According to Ordinal Severity Scale. The distribution of the clinical status according to the ordinal severity scale at baseline, day 7, day 14, day 21 and day 35 is shown for the CCP group and control group according initial randomization, i.e. the 7 patients with crossover to receive CCP on days 15, 17 and 19 remain in the control group.

**Figure 5:**
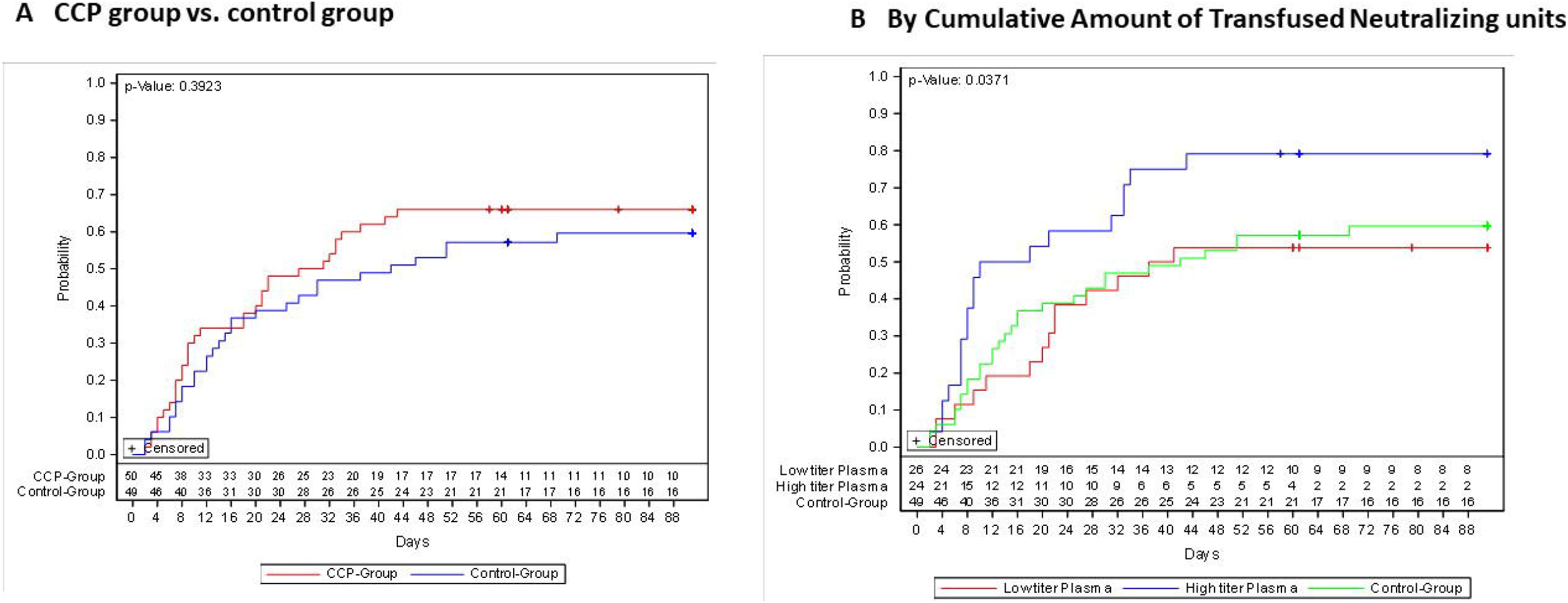
Probability of Discharge from ICU. (A) Probability of discharge from ICU compared in the CCP group (red line) or control group (blue line). Censored patients are indicated by +. p=0.39 (log-rank test). (B) Discharge from ICU compared in the CCP subgroup which received a low cumulative amount of neutralizing units (red), the CCP subgroup which received a high cumulative amount of neutralizing units (blue) and the control group (green line). Censored patients are indicated by +. p=0.04 (log-rank test, high amount group vs. control group).

The secondary time to event outcomes by inflammation markers at baseline are summarized in **Appendix**, Table 4 and Figure 3. For those patients with high inflammation markers the median time to clinical improvement was not reached (**Appendix**, Table 4, Figure 3a). Among patients with low inflammation markers at baseline, the median time to clinical improvement was 19 days (IQR 8-n.r.) in the CCP group and 41 days (IQR 13-n.r.) in the control group. Among patients with high inflammation markers, the time to discharge from hospital was 41 days (IQR 21-n.r.) in the CCP group and was not reached in the control group (**Appendix**, Table 4 and Figure 3b). Among patients with low inflammation markers the median time to hospital discharge was 24 days (IQR 11-n.r.) in the CCP group and 26 days (IQR 12-n.r.) in the control group.

The secondary time to event outcomes by presence of neutralizing antibodies at baseline and by the ventilation status at baseline are summarized in **Appendix** (Table 5 and Table 6).

### Crossover patients

Baseline characteristics of crossover patients are summarized in **Appendix** (Table 7). None of the crossover patients achieved clinical improvement and hospital discharge and all seven patients died (**Appendix**, Figure 4). We compared the crossover patients with a propensity score matched subset of patients from the initial CCP group: patients matched according to ventilation status on day +14 and baseline characteristics (**Appendix**, Table 8) showed no difference in overall survival. In contrast, a subset matched for baseline characteristic only (**Appendix**, Table 9) showed a significant difference in overall survival (**Appendix**, Figure 5).

## Discussion

This trial found no statistically significant difference in the primary outcome, a dichotomous composite outcome of survival without ventilation support at day 21 after treatment with CCP compared to standard treatment in hospitalized patients who required supplemental oxygen, non-invasive or invasive ventilation or ICU treatment. The secondary outcomes time to clinical improvement, time to ICU discharge, time to hospital discharge and case fatality rate were consistently numerically better in the CCP group than in the control group, however not statistically significant.

In two subgroup analyses, we could demonstrate that the primary outcome was better in patients with low inflammation markers at baseline and in patients not requiring invasive ventilation or ECMO at baseline. However, in the subgroup analysis the primary and secondary outcomes did not differ significantly between the treatment groups. A better outcome of patients not requiring invasive ventilation is in line with other reports (1;4).

So far there is no consensus on a minimum antibody titer in CCP – mainly due to a lack of standardization of assays for SARS-CoV-2 neutralizing antibodies. While there is an increasing number of studies comparing results of total binding antibodies (18;23), the comparability of assays for neutralizing antibodies is rather limited. Therefore, it is difficult to compare the neutralizing capacity of CCP products used in the different clinical trials. In addition to direct antibody-mediated virus neutralization other mechanisms of action based on Fc-dependent antibody functions (24) and cellular immunological effects (25) may have implications for therapeutic efficacy of CCP. Also, the total volume of transfused CCP matters. In this trial the median total dose of plasma per patient was 846 ml which is higher than in other published clinical trials which administered a total CCP volume of 200 ml (4), 200-250 ml (16), 250 ml (12), 300 ml (7), 400 (5;8;10;17), 500 ml (1;9;11) and 550 ml (15). Despite this high CCP volume the CAPSID trial failed to demonstrate a significant effect on the primary outcome. However, there was a signal of benefit for CCP in a pre-specified subgroup analysis of patients who were treated with a high amount of neutralizing antibodies. The primary outcome was numerically better and the time to clinical improvement, time to ICU discharge and hospital discharge and overall survival were significantly better (exploratory analysis) in the subgroup which has received high amounts of neutralizing units compared to the control group.

For the findings in this trial three characteristics of the patient population might be of relevance: (i) inclusion of patients with respiratory distress in a broad range from supplemental oxygen to invasive ventilation (the latter subgroup comprising about 34%). (ii) the median interval from onset of symptoms to randomization was 7 days. (iii) the majority of patients already had neutralizing antibodies at baseline. Other trials which also included hospitalized patients requiring ventilation support at least in a proportion of patients also did not report significant differences in clinical improvement or all-cause mortality (4-11). However, in the CAPSID trial, the proportion of patients with life-threatening disease, i.e. requiring invasive ventilation or ECMO, was higher than in most other trials (4-11). This trial included a larger proportion of patients with poor prognosis – based on proportion of patients with invasive ventilation, high inflammation markers and comorbidities which are associated with poor outcome (26). Despite a short median interval of 7 days between onset of symptoms and randomization, a majority of patients had already mounted an immune response and the median PRNT50 titer was 1:160 – even before the transfusion of the first CCP unit. Similar findings were reported from other trials (6;7;11;15) and one trial has even been stopped early due to this observation (7).

Thus, late administration of CCP to patients who already had progressed to severe COVID-19 requiring ventilation support or ICU treatment and who already had developed neutralizing antibodies does not significantly improve all-cause mortality or time to clinical improvement with an effect size which was the basis for the sample size calculation of this trial. In contrast, early treatment with high-titer CCP within 72 hours of onset of mild COVID-19 symptoms reduced to risk of progression to severe respiratory disease by 48% (12). This is in line with our observations in the subgroup analysis by the cumulative amount of transfused neutralizing antibodies and other reports (1;16).

There was no signal that frequency, severity, type and outcome of AEs or SAEs in the CCP group differed from the control group. Rather, there was a tendency of lower number of AEs and lower number of SAEs in the CCP group. Thus, like other studies (4-11;16;27), this trial does not raise concerns that there are new safety issues if plasma is given in the proinflammatory and prothrombotic state of severe COVID-19.

A specific feature of this trial which differs from other trials is the option of crossover of patients from the control group to receive CCP if they present with progressive COVID-19 on day 14. This should address the question whether even late administration of CCP in progressive COVID-19 can improve outcome. Only 7 patients were switched, none had achieved clinical improvement and all have died. The baseline characteristics of the patients who were switched to the CCP arm did not differ from the control group patients who were not switched to receive CCP. The crossover patients were in a poor clinical condition on day 14 – just by the fact that they were eligible for crossover which indicated progressive respiratory disease. The comparison of crossover patients with propensity score matched subgroups from the initial CCP group suggests that the very poor outcome of this small subgroup represents a selection of patients with poor prognosis and unfavorable clinical course irrespective of treatment. Our observation does not support the use of CCP as a last resort in progressive patients.

In conclusion, among hospitalized patients with severe COVID-19, CCP added to standard therapy compared to standard therapy alone did not result in a statistically significant improvement of the primary outcome, i.e. survival free of ventilation support on day 21 and the key secondary outcome time to clinical improvement. The consistent trend for a benefit across all primary and secondary outcomes among patients who have received a higher amount of neutralizing antibodies provides a signal that better outcomes can be achieved by high dose CCP treatment combining both very high titers of neutralizing antibodies with high CCP volumes.

## Supporting information

Clinical Trial Group

## Data Availability

Please send requests regarding availability of anonymized data to.

## Acknowledgement

We are grateful to the members of the Data Safety Monitoring Board for their advice (Prof.Dr.Jan Beyersmann; Prof.Dr.Christian von Heymann; Prof.Dr.Rainer Seitz; Prof.Dr.Carl Kirchmaier), Philipp Schnecko M.Sc., Dr.Christine Windemuth-Kieselbach, Dr.Martin Graeßner and Dr.Gerlinde Schmidtke-Schrezenmeier for expert advice.

We thank all patients who participated in this trial. We thank the clinical research teams, physicians, study nurses and data managers in all clinical trial centers and the team of the CRO Alcedis. Conducting a clinical trial during a pandemic is a challenge. The commitment to take on substantial extra work when health care resources are already fully occupied by daily care for the severely sick patients deserves special recognition!

The clinical trial CAPSID is supported by the Bundesministerium für Gesundheit (“German Federal Ministry of Health”).

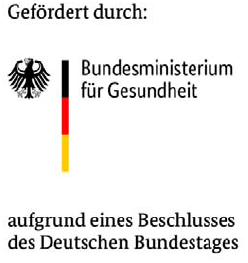

## Appendix

List of members of the CAPSID Clinical Trial Group.

