## Supplementary material for "High Dose Convalescent Plasma in COVID-19: Results from the Randomized Trial CAPSID": Online Supplement

### Appendix: Online Material

#### Supplementary Tables

Table 1: Adverse Events
Table 2: Case Fatality Rate at Days 21, 35 and 60
Table 3: Time to Event for Secondary Outcome by Cumulative Amount of Transfused Neutralizing Units
Table 4: Time to Event for Secondary Outcome by Inflammation Marker
Table 5: Time to Event for Secondary Outcome by presence or absence of neutralizing antibodies at baseline
Table 6: Time to Event for Secondary Outcome by ventilation status at baseline.
Table 7: Baseline Demographics and Clinical Characteristics of patients in the control group with or without crossover due to progressive disease on day 14.
Table 8: Baseline Demographics and Clinical Characteristics of Patients in the Control Group with Crossover and Patients from the CCP Group Matched by Baseline Characteristics.
Table 9: Baseline Demographics and Clinical Characteristics of Patients in the Control Group with Crossover and Patients from the CCP Group Matched by Ventilation Status at Day 14.

#### Supplementary Figures

Figure 1: Distribution of anti-SARS-CoV-2 titer (PRNT50) in transfused CCP
Figure 2a: Probability of first negative SARS-CoV-2 PCR
Figure 2b: Probability of first negative SARS-CoV-2 PCR by Cumulative Amount of Transfused Neutralizing Units
Figure 3a: Probability of Clinical Improvement by Inflammation Markers at Baseline
Figure 3b: Probability of Discharge from Hospital by Inflammation Markers at Baseline
Figure 4: Occurrence of Secondary Outcomes in Control Group Patients with and without Crossover to CCP Treatment.
Figure 5: Probability of Overall Survival of Crossover Patients and Matched Patients from the initial CCP Group.

#### Methods

#### Tables

**Table 1: Adverse Events**

| | CCP group (n=53) | Control Group (n=52) | p-value |
|---|---|---|---|
| Frequency of adverse events, n (%) | | | 0.62 |
| 0 | 11 (20.8) | 9 (17.3) | |
| 1 | 12 (22.6) | 8 (15.4) | |
| 2 | 7 (13.2) | 6 (11.5) | |
| 3 | 11 (20.8) | 7 (13.5) | |
| 4 | 4 ( 7.6) | 6 (11.5) | |
| 5 | 1 ( 1.9) | 5 ( 9.6) | |
| 6 | 3 ( 5.7) | 5 ( 9.6) | |
| ≥7 | 4 ( 7.6) | 6 (11.5) | |
| Worst AE grade, n (%) | | | 0.18 |
| No AE | 11 (20.8) | 9 (17.3) | |
| 1 | 4 ( 7.6) | 0 ( 0.0) | |
| 2 | 9 (17.0) | 6 (11.5) | |
| 3 | 7 (13.2) | 12 (23.1) | |
| 4 | 11 (20.8) | 8 (15.4) | |
| 5 | 11 (20.8) | 17 (32.7) | |
| Outcome, n (%)* | | | |
| Resolved | 32 (60.4) | 36 (69.2) | |
| Ongoing/resolving | 19 (35.9) | 19 (36.5) | |
| Resolved with sequelae | 4 ( 7.6) | 4 ( 7.7) | |
| Death | 11 (20.8) | 17 (32.7) | |
| Unknown | 6 (11.3) | 8 (15.4) | |
| Causal relationship to IMP, n (%)*, ** | | | |
| None | 34 (64.2) | 43 (82.7) | |
| Unlikely | 18 (34.0) | 8 (15.4) | |
| Possible | 3 ( 5.7) | 1 ( 1.9) | |
| Definitive | 0 ( 0.0) | 0 ( 0.0) | |
| Patients with SAE, n (%) | | | |
| No | 31 (58.5) | 27 (51.9) | |
| Yes | 22 (41.5) | 25 (48.1) | |
| Number of SAE, n (%) | | | |
| 1 | 12 (22.6) | 13 (25.0) | |
| 2 | 6 (11.3) | 5 ( 9.6) | |
| 3 | 2 ( 3.8) | 5 ( 9.6) | |
| 4 | 1 ( 1.9) | 2 ( 3.9) | |
| 6 | 1 ( 1.9) | 0 ( 0.0) | |
| Reason for Seriousness, n (%)* | | | |
| Life threatening | 16 (30.2) | 15 (28.9) | |
| Persistent / significant disability | 2 ( 3.8) | 1 ( 1.9) | |
| Resulted in death | 11 (20.8) | 17 (32.7) | |
| Hospitalization / prolongation | 7 (13.2) | 6 (11.5) | |
| Important medical event | 5 ( 9.4) | 6 (11.5) | |

*Multiple answers possible. **Control group patients contain also patients who switched to the plasma arm.

**Table 2: Case Fatality Rate at Days 21, 35 and 60.**

| | CCP-Group (n=53) | Control-Group (n=52) | p-Value | Total |
|---|---|---|---|---|
| **Day 21** | | | 0.79 | |
| Patient alive | 46 (86.8) | 44 (84.6 ) | | 90 (85.7) |
| Patient dead | 7 (13.2) | 8 (15.4) | | 15 (14.3) |
| **Day 35** | | | 0.16 | |
| Patient alive | 45 (84.9) | 38 (73.1) | | 83 (79.1) |
| Patient dead | 8 (15.1) | 14 (26.9) | | 22 (21.0) |
| **Day 60** | | | 0.19 | |
| Patient alive | 42 (79.3) | 35 (67.3) | | 77 (73.3) |
| Patient dead | 11 (20.8) | 17 (32.7) | | 28 (26.7) |

**Table 3: Time to Event for Secondary Outcome by Transfused Neutralizing Units***

| | CCP group (n=53) | | Control Group (n=52) |
|---|---|---|---|
| | Low neutralizing units* (n=28) | High neutralizing units* (n=25) | |
| **Time to clinical improvement, days** | | | |
| **N** | 28 | 25 | 52 |
| **Median (IQR)** | 36 (17-n.r.) | 20 (11-n.r.) | 66 (13-n.r.) |
| **Time to discharge from ICU, days** | | | |
| **N** | 26 | 24 | 49 |
| **Median (IQR)** | 39 (20-n.r.) | 14 ( 7-39) | 42 (12-n.r.) |
| **Time to discharge from hospital, days** | | | |
| **N** | 28 | 25 | 52 |
| **Median (IQR)** | 39 (21-n.r.) | 21 (13-43) | 51 (20-n.r) |
| **Time to first negative SARS-CoV-2 PRC, days** | | | |
| **N** | 28 | 25 | 52 |
| **Median (IQR)** | 14 ( 5-19) | 5 ( 3-15) | 8 ( 5-21) |

*see Methods section for definition of neutralizing units. CCP group was divided by the cumulative amount of neutralizing units per patient (all 3 CCP transfusions) in a low neutralizing unit group (≤ median) and a high neutralizing unit group (> median). n.r., not reached.

**Table 4: Time to Event for Secondary Outcome by Inflammation Markers***

| | High Inflammation Markers (n=51)** | | Low inflammation Markers (n=48)** | |
|---|---|---|---|---|
| | CCP Group | Control Group | CCP Group | Control Group |
| **Time to Clinical Improvement (days)** | | | | |
| **N** | 25 | 26 | 24 | 24 |
| **Median (IQR)** | n.r. (21-n.r.) | n.r. (11-n.r.) | 19 (8-n.r.) | 41 (13-n.r.) |
| **Time to Discharge from ICU (days)** | | | | |
| **N** | 24 | 26 | 22 | 21 |
| **Median (IQR)** | 42 (20-n.r.) | n.r. (12-n.r.) | 13 ( 6-37) | 20 (12-69) |
| **Time to Discharge from hospital (days)** | | | | |
| **N** | 25 | 26 | 24 | 24 |
| **Median (IQR)** | 41 (21-n.r.) | n.r. (30-n.r.) | 24 ( 11-n.r.) | 26(12-n.r.) |
| **Time to first negative SARS-CoV-2 PCR** | | | | |
| **N** | 25 | 26 | 24 | 24 |
| **Median (IQR)** | 14 ( 5-18) | 15 ( 5-n.r.) | 5 ( 3-18) | 6 ( 5-18) |

*see Methods section for definition of high / low inflammation markers. The patient group was divided in a low inflammation marker group and a high neutralizing unit group. In each of these groups the primary outcome for patients in the CCP group and the control group was compared. ** Six patients with either missing data on inflammation markers (n=1) or intermediate inflammation markers (n=5) are not included in this table. n.r., not reached.

**Table 5: Time to Event for Secondary Outcome by Presence or Absence of Neutralizing Antibodies at Baseline***

| | anti-SARS-CoV-2 neutralizing antibodies at baseline: positive (n=75)** | | anti-SARS-CoV-2 neutralizing antibodies at baseline: negative (n=20)** | |
|---|---|---|---|---|
| | CCP Group | Control Group | CCP Group | Control Group |
| Time to Clinical Improvement (days) | | | | |
| N | 37 | 38 | 10 | 10 |
| Median (IQR) | 32 (15-n.r.) | n.r. (14-n.r.) | 27 (19-n.r.) | 32( 9-n.r.) |
| Time to Discharge from ICU (days) | | | | |
| N | 35 | 36 | 9 | 9 |
| Median (IQR) | 27 ( 9-n.r.) | 40 (11-n.r.) | 33 ( 7-n.r.) | 25 (15-n.r.) |
| Time to Discharge from hospital (days) | | | | |
| N | 37 | 38 | 10 | 10 |
| Median (IQR) | 27 (16-n.r.) | 37 (14-n.r.) | n.r. (14-n.r.) | n.r. ( 29-n.r.) |
| Time to first negative SARS-CoV-2 PCR | | | | |
| N | 37 | 38 | 10 | 10 |
| Median (IQR) | 7 ( 4-15) | 7 ( 5-22) | 16 (4-19) | 15 ( 7-19) |

* presence of anti-SARS-CoV-2 neutralizing antibodies PRNT50 ≥ 1:20 at baseline.

** Ten patients with missing data on neutralizing antibodies (PRNT50) at baseline are not included in this table.

**Table 6: Time to Event for Secondary Outcome by Ventilation Status at Baseline.**

| | Patients without invasive ventilation / ECMO (n=69) | | Patients wit invasive ventilation / ECMO (n=36) | |
|---|---|---|---|---|
| | CCP Group | Control Group | CCP Group | Control Group |
| **Time to Clinical Improvement (days)** | | | | |
| **N** | 37 | 32 | 16 | 20 |
| **Median (IQR)** | 21 (15-n.r.) | n.r. (15-n.r.) | 33 (12-n.r.) | 66 (13-n.r.) |
| **Time to Discharge from ICU (days)** | | | | |
| **N** | 34 | 29 | 16 | 20 |
| **Median (IQR)** | 16 ( 7-n.r.) | 16 ( 8-n.r.) | n.r. (32-n.r.) | n.r. (29-n.r.) |
| **Time to Discharge from Hospital (days)** | | | | |
| **n** | 37 | 32 | 16 | 20 |
| **Median (IQR)** | 21 (13-n.r.) | 34 (12-n.r.) | n.r. (32-n.r.) | n.r. (29-n.r.) |
| **Time to first negative SARS-CoV-2 PCR** | | | | |
| **n** | 37 | 32 | 16 | 20 |
| **Median (IQR)** | 7 ( 4-18) | 8 ( 5-26) | 7 ( 5-16) | 11 ( 5-19) |

n.r., not reached.

**Table 7: Baseline Demographics and Clinical Characteristics of Patients in the Control Group with or without Crossover Due to Progressive Disease on Day 14.**

| | Crossover (n=7) | No crossover (n=45) | p-value |
|---|---|---|---|
| **Demographic and clinical characteristics** | | | |
| **Median age, years (IQR)** | 62 (53-63) | 62 (55-67) | 0.62 |
| **Gender, no (%)** | | | 1.00 |
| Female | 2 (28.6) | 15 (33.3) | |
| Male | 5 (71.4) | 30 (67.3) | |
| **Body Mass Index, kg/$m^2$ (IQR)** | 29.0 (23.2-30.4) | 29.4 (26.1-32.00) | 0.29 |
| **Coexisting Diseases, n (%)** | | | |
| no other disease | 1 (14.3) | 3 ( 6.7) | |
| BMI >30 kg/m2 | 5 (71.4) | 24 (53.3) | |
| Hypertension | 3 (42.9) | 25 (55.6) | |
| Diabetes | 1 (14.3) | 14 (31.1) | |
| COPD, Asthma, other pulmonary disease | 1 (14.3) | 8 (17.8)) | |
| Thromboembolic disease | 1 (14.3) | 2 ( 4.4) | |
| solid tumor | 1 (14.3) | 2 ( 4.4) | |
| other | 4 (57.1) | 31 (68.9) | |
| **Point Scale at Study entry, n (%)** | | | 0.79 |
| 3 | 0 ( 0.0) | 3 ( 6.7) | |
| 4 | 0 ( 0.0) | 8 (17.8) | |
| 5 | 4 (57.1) | 17 (37.8) | |
| 6 | 0 ( 0.0) | 3 ( 6.7) | |
| 7 | 3 (42.9) | 14 (31.1) | |
| **Median time from symptom onset of the SARS-CoV-2 infection to randomization, days (IQR)** | 10 (7-10) | 7 (5-11) | 0.88 |
| **Median time from hospitalization to randomization, days (IQR)§** | 3 (1-8) | 2 (1-5) | 0.77 |

| | | | |
|---|---|---|---|
| **SARS-CoV-2 status at baseline** | | | |
| **Result of SARS-CoV-2 PCR nasopharyngeal swab, n (%)** | | | 1.0 |
| **positive** | 7 (100.0) | 43 (95.6) | |
| **negative** | 0 ( 0.0) | 1 ( 2.2) | |
| **missing** | 0 ( 0.0) | 1 ( 2.2) | |
| **Humoral immune response at baseline** | | | |
| **SARS-CoV-2 IgA present, n (%)** | | | 0.82 |
| **Yes** | 6 (85.7) | 32 (71.1) | |
| **No** | 1 (14.3) | 6 (13.3) | |
| **Missing** | 0 ( 0.0) | 7 (15.6) | |
| **SARS-CoV-2 IgG present, n (%)** | | | 0.85 |
| **Yes** | 5 (71.4) | 25 (55.6) | |
| **No** | 2 (28.6) | 13 (28.9) | |
| **Missing** | 0 (0) | 7 (15.5) | |
| **Neutralizing antibodies (based on PRNT50 titer ≥20) present** | | | 0.63 |
| **no** | 2 (28.6) | 8 (17.8) | |
| **yes** | 5 (71.4) | 33 (73.3) | |
| **Missing** | 0 ( 0.0) | 4 ( 8.9) | |

| | | |
|---|---|---|
| **Laboratory values at baseline** | | |
| **Inflammation Markers** | | |
| **Ferritin [µg/L], median (IQR)** | 1412 (1015-1467) | 1096 (639-2496) |
| **CRP [mg/L], median (IQR)** | 157 (102-227) | 126 (66-194) |
| **IL-6 [pg/ml], median (IQR)** | 142 (44-253) | 35 (17-104) |
| **LDH [U/L], median (IQR)** | 631 (477-789) | 481 (362-657) |

**Table 8: Baseline Demographics and Clinical Characteristics of Patients in the Control Group with Crossover and Patients from the CCP Group Matched by Baseline Characteristics.**

| | Crossover (n=7) | CCP (n=14) | p-value |
|---|---|---|---|
| **Demographic and clinical characteristics** | | | |
| **Median age, years (IQR)** | 62 (53-63) | 57 (50-64) | 0.66 |
| **Gender, no (%)** | | | 0.57 |
| **Female** | 2 (28.6) | 2 (85.7) | |
| **Male** | 5 (71.4) | 12 (14.3) | |
| **Body Mass Index, kg/m² (IQR)** | 29.0 (23.2-30.4) | 30.8 (27.5-93.4) | 0.09 |
| **Coexisting Diseases, n (%)** | | | |
| **no other disease** | 1 (14.3) | 1 (7.1) | |
| **BMI >30 kg/m2** | 5 (71.4) | 6 (42.9) | |
| **Hypertension** | 3 (42.9) | 9 (64.3) | |
| **Cardiovascular disease** | 0 (0.0) | 3 (21.4) | |
| **Diabetes** | 1 (14.3) | 6 (42.9) | |
| **COPD, Asthma, other pulmonary disease** | 1 (14.3) | 3 (21.2)) | |
| **Thromboembolic disease** | 1 (14.3) | 0 (0) | |
| **solid tumor** | 1 (14.3) | 0 (0) | |
| **other** | 5 (71.4) | 7 (50.0) | |
| **Point Scale at Study entry, n (%)** | | | 0.92 |
| **3** | 0 ( 0.0) | 1 ( 7.1) | |
| **4** | 0 ( 0.0) | 2 (14.3) | |
| **5** | 4 (57.1) | 5 (35.7) | |
| **6** | 0 ( 0.0) | 1 ( 7.1) | |
| **7** | 3 (42.9) | 5 (36.7) | |
| **Respiratory support at baseline** | | | 0.96 |
| **No respiratory support** | 0 ( 0) | 1 ( 7.1) | |
| **$O_2$ by nasal cannula** | 0 ( 0) | 2 (14.3) | |
| **High flow $O_2$** | 2 (28.6) | 2 (14.3) | |
| **Non-invasive ventilation** | 2 (28.6) | 3 (21.4) | |
| **Invasive ventilation** | 3 (42.9) | 5 (35.7) | |
| **ECMO** | 0 ( 0) | 1 (7.1) | |
| **Median time from symptom onset of the SARS-CoV-2 infection to randomization, days (IQR)** | 10 (7-10) | 7 (3-8) | 0.17 |
| **Median time from hospitalization to randomization, days (IQR)§** | 3 (1-8) | 1 (1-2) | 0.11 |
| **Transfused neutralizing units Median (IQR)** | 2952 (1688-3348) | 5988 (3328-6544) | |

| Humoral immune response at baseline | | | |
|---|---|---|---|
| SARS-CoV-2 IgA present, n (%) | | | 1.00 |
| Yes | 6 (85.7) | 10 (71.4) | |
| No | 1 (14.3) | 3 (21.4) | |
| Missing | 0 ( 0) | 1 ( 7.1) | |
| SARS-CoV-2 IgG present, n (%) | | | 0.44 |
| Yes | 5 (71.4) | 5 (35.7) | |
| No | 2 (28.6) | 8 (57.1) | |
| Missing | 0 ( 0.0) | 1 ( 7.1) | |
| Neutralizing antibodies (based on PRNT50 titer ≥20) present | | | 1.00 |
| no | 2 (28.6) | 4 (28.6) | |
| yes | 5 (71.4) | 10 (71.4) | |

| Laboratory values at baseline | | |
|---|---|---|
| Inflammation Markers | | |
| Ferritin [µg/L], median (IQR) | 1412 (1015-1467) | 1109 (407-1691) |
| CRP [mg/L], median (IQR) | 157 (102-227) | 229 (76-284) |
| IL-6 [pg/ml], median (IQR) | 142 (44-253) | 94 (43-154) |
| LDH [U/L], median (IQR) | 631 (477-789) | 463 (426-571) |

**Table 9: Baseline Demographics and Clinical Characteristics of Patients in the Control Group with Crossover and Patients from the CCP Group Matched by Ventilation Status at Day 14.**

| | Crossover (n=6) | CCP (n=6) | p-value |
|---|---|---|---|
| **Demographic and clinical characteristics** | | | |
| Median age, years (IQR) | 62 (53-63) | 64 (59-68) | 0.29 |
| Gender, no (%) | | | 1.00 |
| Female | 1 (16.7) | 0 ( 0) |  |
| Male | 5 (83.3) | 6 (100) | |
| Body Mass Index, kg/$m^2$ (IQR) | 27.7 (23.2-29.1) | 27.8 (27.8-30.4) | 0.20 |
| Coexisting Diseases, n (%) | | | |
| no other disease | 1 (16.7) | 0 ( 0) |  |
| BMI >30 kg/m2 | 5 (83.3) | 4 (66.7) | |
| Hypertension | 2 (33.3) | 5 (83.3) | |
| Cardiovascular disease | 0 ( 0.0) | 2 (33.3) | |
| Diabetes | 0 ( 0) | 2 (33.3) |  |
| COPD, Asthma, other pulmonary disease | 1 (16.7) | 3 (50.0) | |
| Thromboembolic disease | 1 (16.7) | 0 ( 0) |  |
| solid tumor | 1 (16.7) | 0 ( 0) |  |
| other | 4 (57.1) | 4 (57.1) | |
| Point Scale at day 14, n (%) | | | 0.18 |
| 6 | 0 ( 0) | 3 (50.0) |  |
| 7 | 6 (100) | 3 (50.0) | |
| Respiratory support at day 14, n (%) | | | 0.24 |
| Invasive ventilation | 1 (16.7) | 4 (66.7) | |
| ECMO | 5 (83.3) | 2 (33.3) | |
| Median time from symptom onset of the SARS-CoV-2 infection to randomization, days (IQR) | 10 (8-10) | 5 (2-9) | 0.18 |
| Median time from hospitalization to randomization, days (IQR)[§0] | 4 (2-8) | 2 (1-2) | 0.26 |
| Transfused neutralizing units Median (IQR) | 3092 (2362-3348) | 4844 (3384-6336) | |

| Humoral immune response at baseline | | | |
|---|---|---|---|
| **SARS-CoV-2 IgA present, n (%)** | | | 1.00 |
| Yes | 6 (100) | 6 (100) | |
| No | 0 ( 0) | 0 ( 0) | |
| **SARS-CoV-2 IgG present, n (%)** | | | 1.00 |
| Yes | 5 (83.3) | 5 (83.3) | |
| No | 1 (16.7) | 1 (16.7) | |
| **Neutralizing antibodies (based on PRNT50 titer ≥20) present** | | | 1.00 |
| no | 1 (16.7) | 0 ( 0) | |
| yes | 5 (83.3) | 6 (100) | |

| Laboratory values at baseline | | |
|---|---|---|
| **Inflammation Markers** | | |
| **Ferritin [µg/L], median (IQR)** | 1432 (1237-1467) | 1002 (817-1433) |
| **CRP [mg/L], median (IQR)** | 154 (102-163) | 218 (81-300) |
| **IL-6 [pg/ml], median (IQR)** | 109 (44-155) | 82 (74-103) |
| **LDH [U/L], median (IQR)** | 559 (477-685) | 515 (456-537) |

#### Figures

##### Figure 1: Distribution of anti-SARS-CoV-2 Titer (PRNT50) in Transfused CCP

The graphs show the relative proportion of patients receiving a CCP with the titer level as indicated on the x-axis for 1., 2. and 3.transfusion of CCP. Titer levels indicate 50% inhibition in the PRNT assay.

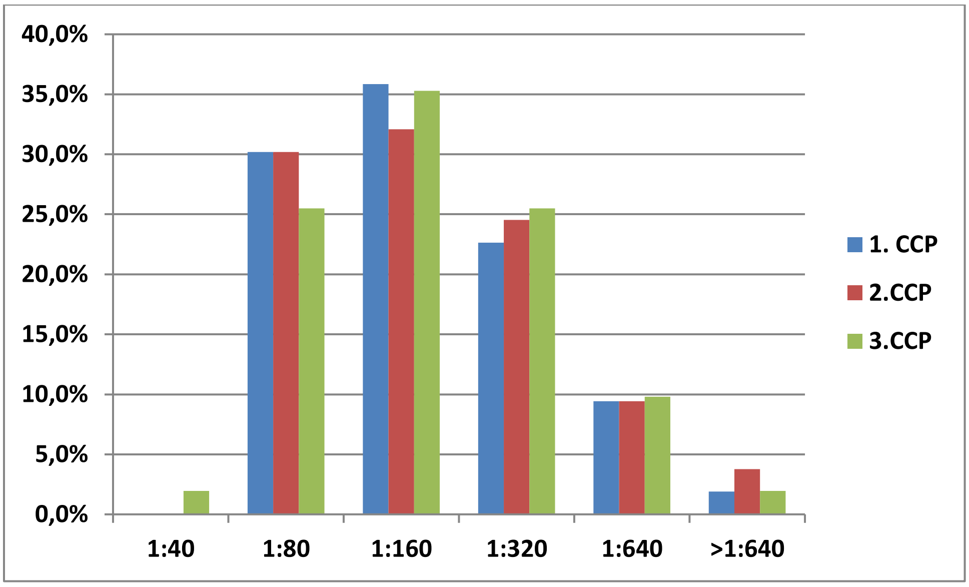

**Figure 2a: Probability of First Negative SARS-CoV-2 PCR**

Kaplan-Meier estimate of probability of first negative SARS-CoV-2 PCR from nasopharyngeal specimen. Censored patients are indicated by +. p=0.38 (log-rank test).

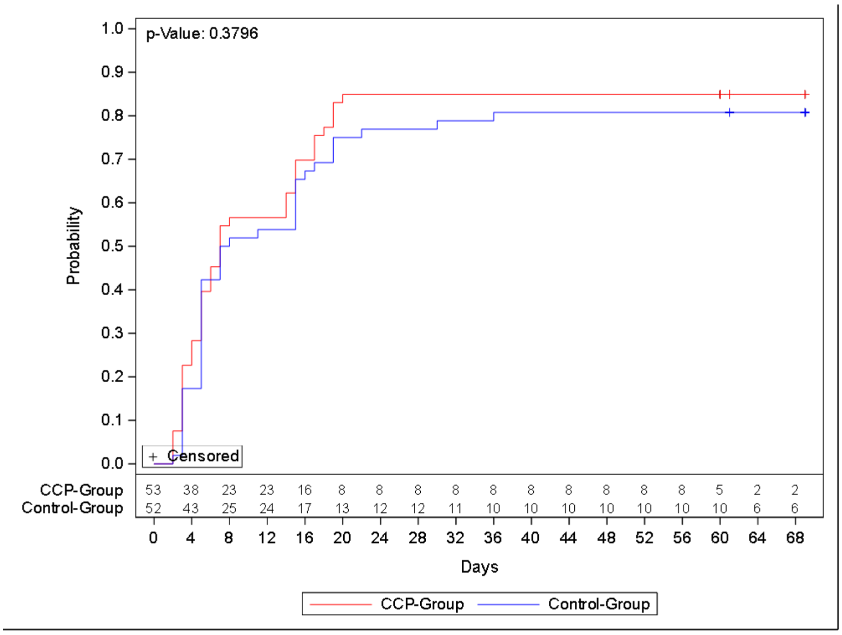

**Figure 2b: Probability of First Negative SARS-CoV-2 PCR by Cumulative Amount of Tranfused Neutralizing Units**

Kaplan-Meier estimate of time to first negative SARS-CoV-2 PCR from nasopharyngeal specimens compared in the CCP subgroup which received a low cumulative amount of neutralizing units (red), the CCP subgroup which received a high cumulative amount of neutralizing units (blue) and the control group (green line). Censored patients are indicated by +. p=0.07 (log-rank test, high amount vs. control group).

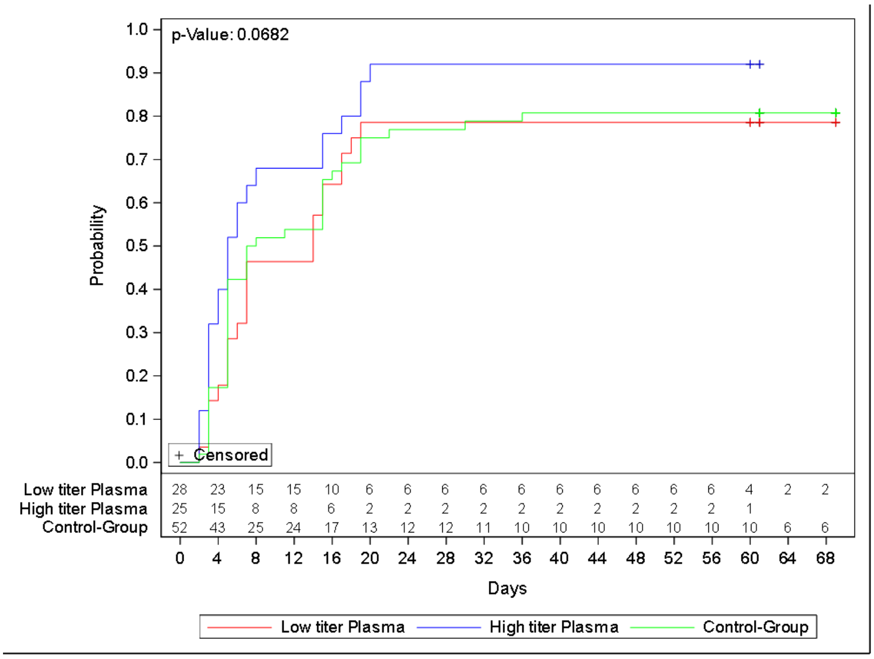

**Figure 3a: Probability of Clinical Improvement by Inflammation Markers at Baseline**

Kaplan-Meier cumulative estimate of probability of clinical improvement compared in the CCP group with low (purble line) and high inflammation marker (red) and the control group with low (turquoise line) and high inflammation markers (blue line) at baseline. Censored patients are indicated by +. p=0.02 (log-rank test).

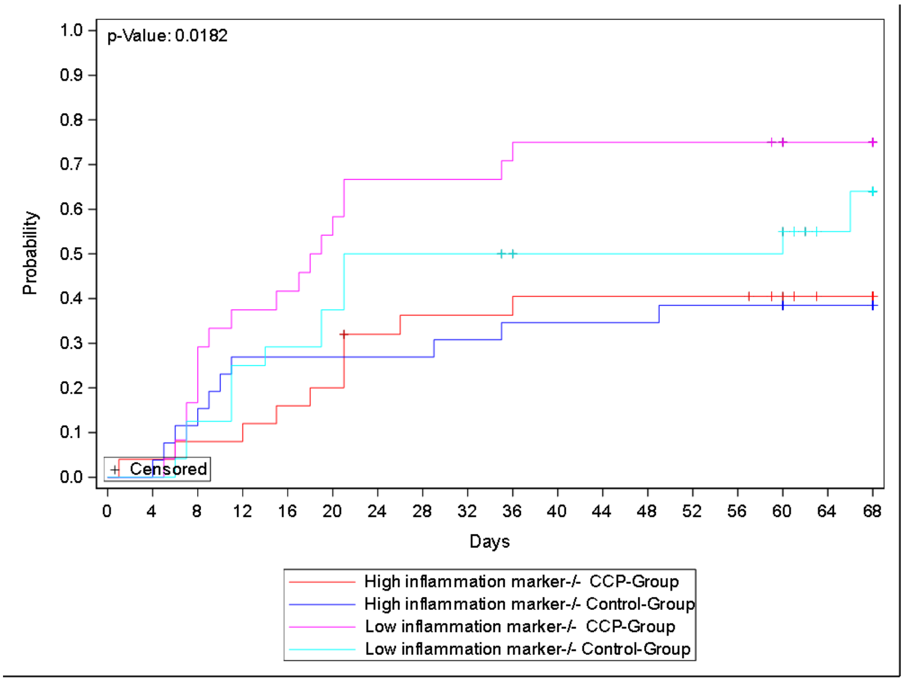

**Figure 3b: Probability of Discharge from Hospital by Inflammation markers at Baseline**

Kaplan-Meier cumulative estimate of probability of discharge from hospital compared in the CCP group with low (purble line) and high inflammation marker (red) and the control group with low (turquoise line) and high inflammation markers (blue line) at baseline. Censored patients are indicated by +. p=0.02 (log-rank test).

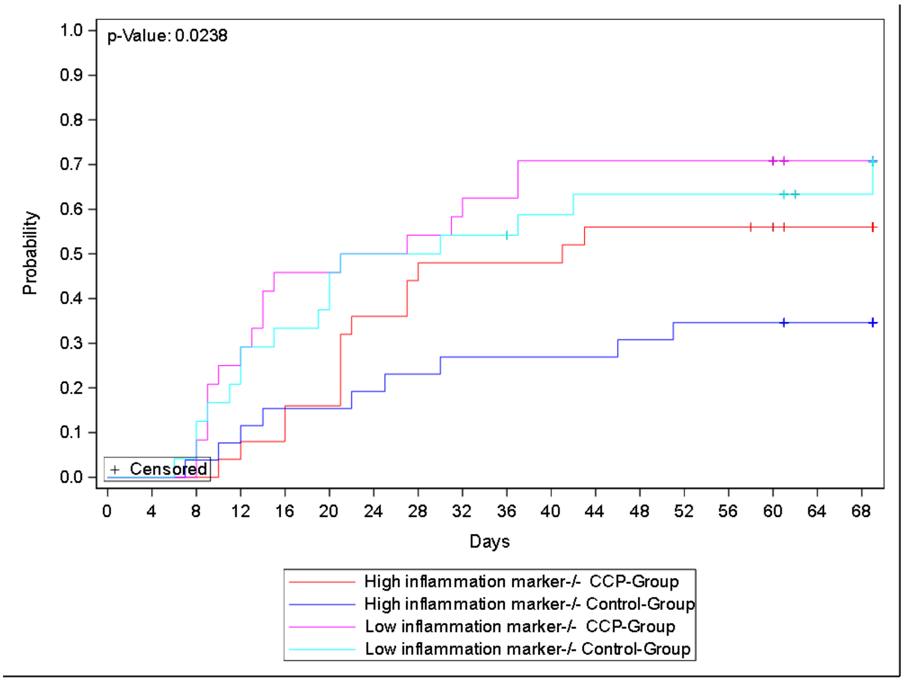

**Figure 4: Occurrence of Secondary Outcomes in Control Group Patients with or without Crossover to CCP Treatment**

**A Probability of clinical improvement**

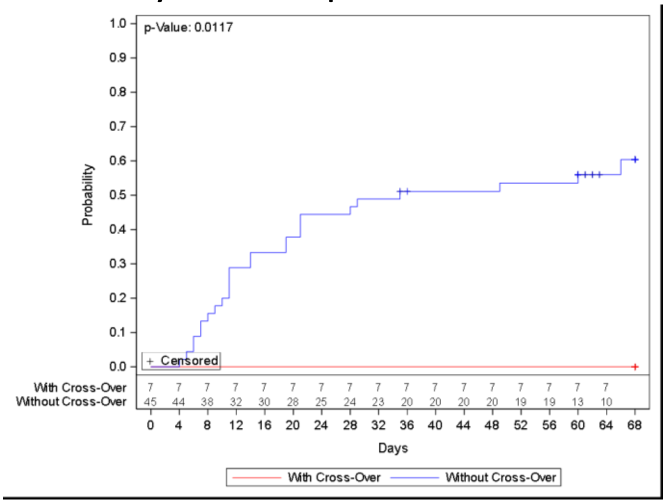

Kaplan-Meier cumulative estimate of probability of

(A) clinical improvement compared in the control group with crossover (red) and without crossover (blue) due to progressive disease on day +14 (CCP treatment on days 15, 17 and 19). Censored patients are indicated by +. P=0.01 (log-rank test).

**B Probability of hospital discharge**

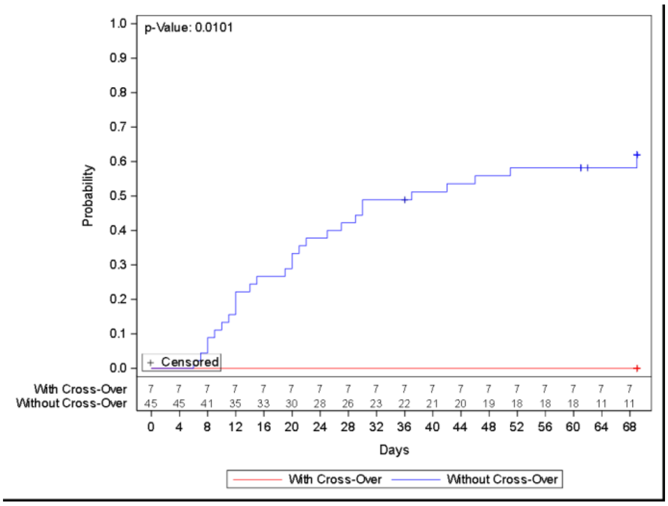

(B) discharge from hospital compared in the control group with crossover (red) and without crossover (blue)) due to progressive disease on day +14 (CCP treatment on days 15, 17 and 19). Censonred patients are indicated by +. P=0.01 (log-rank test).

**C Probability of overall survival**

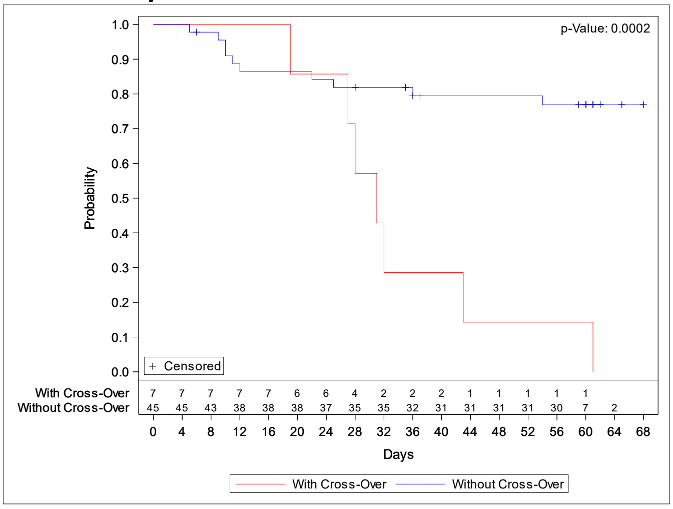

(C) Overall survival in the control group with crossover (red) and without crossover (blue) due to progressive disease on day +14 (CCP treatment on days 15, 17 and 19). Censonred patients are indicated by +. P<0.001 (log-rank test).

**Figure 5: Probability of Overall Survival of Crossover Patients and Matched Patients from the initial CCP Group.**

**A: Crossover patients and CCP group patients matched based on baseline characteristics.**

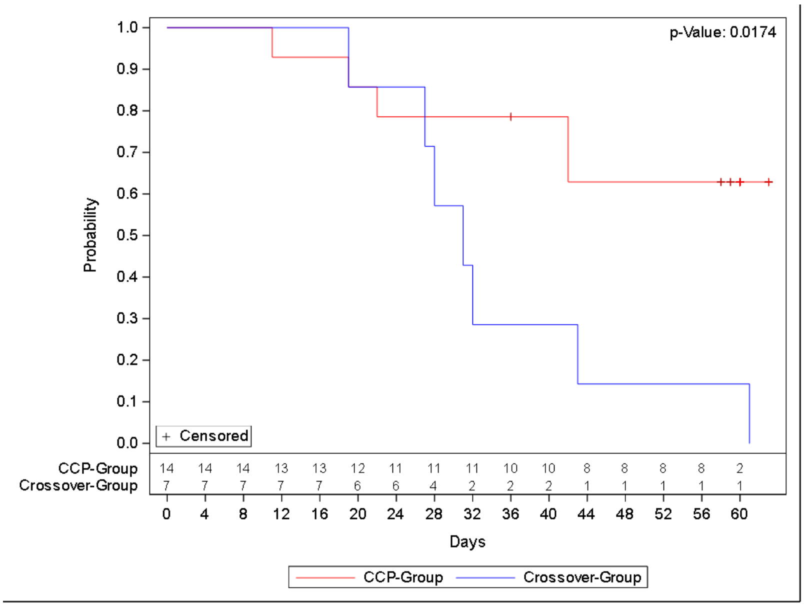

Overall survival in patients with crossover (blue line) treated with CCP on day +15, +17 and +19 (n=7) and matched patients treated with CCP on day +1, +3 and +5 (red line)(n=14). The 14 patients of the plasma group were matched according to baseline characteristics. Censored patients are indicated by +. p=0.017 (log-rank test). Variables for propensity score matching are described in Methods in the Appendix.

**B: Crossover patients and CCP group patients matched based on baseline characteristics and ventilation status on day 14**

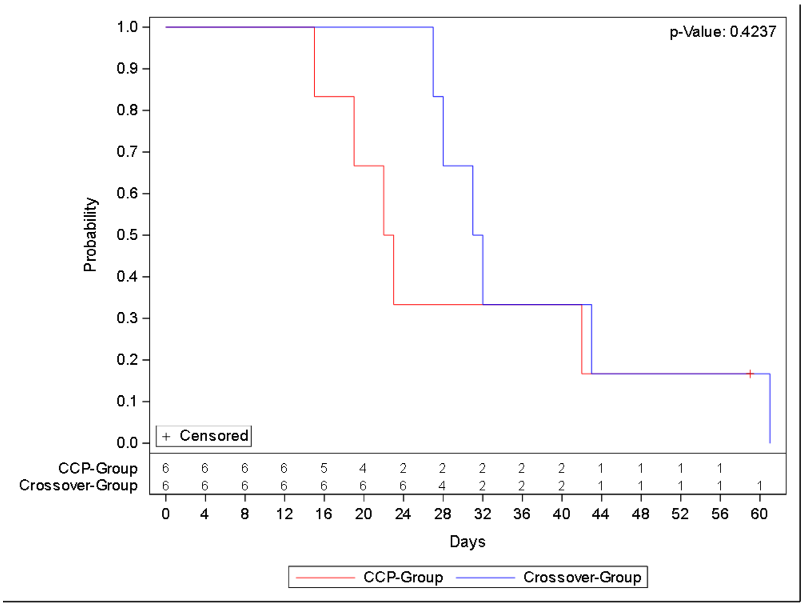

Overall survival in patients with crossover (blue line) treated with CCP on day +15, +17 and +19 (blue line)(n=6) and matched patients treated with CCP on day +1, +3 and +5 (red line)(n=6). The 6 patients of the plasma group were matched according to ventilation status on day 14 and baseline characteristics. Censored patients are indicated by +. p=0.424 (log-rank test). Variables for propensity score matching are described in Methods in the Appendix.

### Methods

#### Additional Information on Patients and Trial Design

##### Exclusion criteria

Exclusion criteria were: (1) accompanying diseases other than COVID-19 with an expected survival time of less than 12 months; (2) previous treatment with any SARS-CoV-2-convalescent plasma; (3) in the opinion of the clinical team, progression to death is imminent and inevitable within the next 48 hours, irrespective of the provision of treatment; (4) Interval > 72 hours since start of mechanical ventilation; (5) not considered eligible for extracorporeal oxygenation support; (6) chronic obstructive lung disease (COPD), stage 4; (7) lung fibrosis with usual interstitial pneumonia pattern in CT and severe emphysema; (8) chronic heart failure NYHA ≥ 3 and/or pre-existing reduction of left ventricular ejection fraction to ≤ 30%; (9) shock of any type requiring ≥ 0.5 µg/kg/min noradrenaline (or equivalent) or requiring more than two types of vasopressor medication for more than 8 hours; (10) liver cirrhosis Child C; (11) liver failure: bilirubin > 5 x upper limit of normal (ULN) and elevation of ALT /AST (at least one >10 x ULN); (12) any history of adverse reactions to plasma proteins; (13) known deficiency of immunoglobulin A; (14) pregnancy; (15) breastfeeding women; (16) volume overload until sufficiently treated and (17) participation in another clinical trial with an investigational medicinal product

##### Crossover from Control Group to CCP treatment

Clinical condition in all patients was evaluated on day 14. In case of progressive COVID-19 on day 14 compared to baseline, patients in the control group could be switched to treatment with CCP. A patient switching from the control group to CCP because of progressive COVID-19 on day 14 was considered as failure of the primary outcome. Criteria of progress for the crossover decision were as follows: (1) in patients not requiring invasive ventilation or ECMO at baseline: start of invasive ventilation support or ECMO in the interval from randomization to day 14; (2) In patients requiring invasive ventilation already at baseline: deterioration of ARDS according to the Berlin classification [1]: (a) progression to moderate or severe ARDS if ARDS was mild at baseline; (b) progression to severe ARDS if moderate at baseline and (c) start of extracorporeal oxygenation in the interval from randomization to day 14. Crossover decisions were confirmed by an independent expert who was experienced in treatment of ARDS and who was not part of the study teams. Seven patients randomized to the control group crossed over to receive CCP after assessment on day 14.

In order to investigate the crossover patients in more detail two subsets of the initial CCP cohort were created. The baseline matched subset of ITT defined by propensity score matching patients from the initial CCP-Group to patients from the Control-Group that had a crossover to CCP by the following variables (Terms in Brackets are possible values)

- age at registration (continuous variable)
- PRNT50-Value at Baseline (Positive; Negative)
- inflammation markers at baseline (High; Intermediate; Low; Missing)
- sex (Male; Female)
- ventilation support at Baseline (ECMO or invasive ventilation; No ECMO or invasive ventilation)
- transfused neutralizing units (>Median over all patients; ≤Median over all patients)

And a second subset characterized by day 14 matched set(D14MS): Subset of ITT defined by propensity score matching patients from the initial CCP-Group to patients from the control-group that had a crossover to CCP by the following variables (terms in brackets are possible values)

- age at registration (continuous variable)
- PRNT50-Value at Baseline (Positive; Negative)
- Inflammation markers at baseline (High; Intermediate; Low; Missing)
- sex (Male; Female)
- ventilation support at Day 14 (ECMO or invasive ventilation; no ECMO or invasive ventilation)
- transfused neutralizing units (>median over all patients; ≤median over all patients)

Propensity score matching was performed by using the SAS procedure PROC PSMATCH with default settings using optimal matching to identify matched patients.

#### Procurement of Convalescent Plasma

Patients who had recovered from SARS-CoV-2 infection were recruited as donors. CCP was collected by plasmapheresis with a median collection volume of 850 ml. The target volume of the CCP units was 250 to 325 ml. The majority of donors had a mild or moderate course of COVID-19. Titers of neutralizing antibodies were measured by a PRNT. A series of 144 donors (41% females, 59% males; median age 40 years) underwent 319 plasmapheresis procedures providing a median collection volume of 850 ml and a mean number of 2.7 therapeutic units per plasmapheresis. The target volume of the CCP units was 250 to 325 ml. The majority of donors had a mild or moderate course of COVID-19. The detailed criteria for acceptance of recovered patients as plasma donors and the correlation of antibody titers with host factors and evolution of neutralizing antibody titers over time in repeat donors were reported elsewhere [2] . Only CCP units with a neutralizing antibody titer of at least 1:20 were accepted. The allocation of CCP to a recipient was based on the following criteria – provided availability: ABO-identical transfusions, all three CCP units for a patient from one donor. If availability of CCP did not allow transfusing ABO-identical plasma, also minor compatible units were used.

#### Definitions of Secondary Outcomes

Case fatality rate at day 21, 35 and 60 was calculated by the number of dead patients divided by the total number of cases at the corresponding point in time. Survival time was time from randomization to death in days. Patients not known to have died were censored at last follow up. Duration of ventilation support was the sum of a patient's single duration of ventilation support and/or ECMO episodes. An episode is defined as the first documentation of a need for non-invasive ventilation or invasive ventilation until the next documentation indicating that no ventilation support (mechanical ventilation, continuous positive airway pressure ventilation, high-flow oxygen, non-invasive ventilation) is needed anymore. Supplemental oxygen (by mask/nasal prongs) in the weaning period was not considered as ventilation support for this analysis. Length of stay in ICU was defined as interval from date of randomization to the first date of discharge from the ICU in days. Patients for whom no discharge from ICU was documented were censored at last follow up. Patients who never entered the ICU were not considered. Length of stay in hospital was defined as interval from date of randomization to the date of hospital discharge (or death) in days. Patients for whom no hospital discharge was documented were censored at last contact. Time until negative SARS-CoV-2 PCR was defined as interval from randomization to first negative SARS-CoV2-PCR. Patients not known to have a negative result were censored at last follow up.

The key secondary outcome time to clinical improvement was defined as an increase by at least two points on the ordinal severity scale. Patients without documented improvement were censored at last follow up.The scale was defined as follows: 0, no clinical or virological evidence of infection; 1, ambulatory without limitation of activities; 2, ambulatory with limitation of activities; 3, hospitalized without oxygen therapy; 4, hospitalized with supplemental oxygen by mask or nasal prongs; 5, hospitalized, non-invasive ventilation or high-flow oxygen; 6, hospitalized, intubation and mechanical ventilation; 7, hospitalized, ventilation and additional organ support (vasopressors, renal replacement therapy or ECMO); 8, death

#### Concomitant Treatment

Patients in both groups received other anti-viral treatment and/or supportive treatment according to institutional standard procedures for patients with severe infection with respiratory viruses. Centers were advised to adhere to current recommendations for treatment of severe COVID-19 published by medical societies. Prior and concomitant medication was coded into certain pre-specified categories (corticosteroids, anticoagulation, tocilizumab, NSAIDS, antiviral drugs, others). Medication was evaluated as concomitant medication when the medication was started after a patient's baseline date.

#### Plaque reduction neutralization test (PRNT) for SARS-CoV-2

Plaque reduction neutralization tests for SARS-CoV-2 were performed as previously described [3-5]. Briefly, VeroE6 cells ($3.25x10^5$ cell/ml) were seeded in 24-well plates and incubated overnight. Prior to PRNT, patient sera were heat-inactivated at 56°C for 30 minutes. For each dilution step (duplicate), patient sera were diluted in 220 µl OptiPro and mixed 1:1 with 220 µl virus solution containing 100 plaque forming units. The 440 µl serum-virus solution was gently vortexed and incubated at 37°C for 1 hour. Each 24-well was incubated with 200 µl serum-virus solution. After 1 hour at 37°C supernatants were discarded, and cells were supplemented with 1.2% Avicel solution in DMEM. After 3 days at 37°C, supernatants were removed and the 24-well plates were fixed and inactivated using a 6% formaldehyde/PBS solution and stained with crystal violet as described (11). Serum dilutions with a plaque reduction of 50% (PRNT50) and 90% (PRNT90) are referred to as titers. Unless stated otherwise, cut off titers were set at < 1:20.

#### Enzyme-linked immunosorbent assay (Euroimmun)

The Euroimmun anti-SARS-CoV-2 assay is a classical enzyme-linked immunosorbent assay (ELISA) for the detection of IgG and IgA to the S1 domain of the SARS-COV-2 spike (S) protein. The assay was performed manually according to the manufacturer´s instructions as previously described [3]. Results are expressed as optical density (OD) ratios, which were calculated based on the sample and calibrator OD values. For all analytes, a ratio < 0.8 was considered to be non-reactive or negative. An OD-ratio of ≥ 1.1 was considered to be positive.

#### Statistical analysis

Sample size calculation was based on an alpha error of 0.05, a power of 0.8, a two-sided comparison and an expected improvement of the primary outcome from 40% (control) to 70% (CCP ). This resulted in a patient number of 48 patients per arm calculated by means of a Fisher´s exact test. Assuming a dropout rate of 10%, the overall number per arm was 53 patients. Sample size calculation was performed with G*Power version 3.1.9.4.

Baseline values of C-reactive protein (CRP), interleukin-6 (IL-6) and ferritin at baseline were compared to their respective median within the total ITT population and patients were allocated by the following rules: (1) at least two non-missing values of inflammation markers below or equal to the median: “Low inflammation markers”; (2) at least two non-missing values above the median : “High inflammation markers”; (3) only two of the three inflammation markers available and one below and one above median: “Intermediate inflammation markers”; (4) all values are missing: “Missing”.

Since the primary outcome is the only confirmatory outcome for this study, an adjustment of the type 1 error due to multiple testing is not required. All other p-values are fully explorative. Secondary outcomes were analyzed using a Kaplan-Meier estimation approach. Patients that died during observation without reaching the secondary outcome will be censored as if they reached the end of observation to account for the competing risk setting. Pre-defined subgroup analyses compared outcome measures in patients with low or high amounts of neutralizing units transfused (cumulative neutralizing units of all transfused CCP products equal or below the median or above the median) and in subgroups with low or high inflammation and coagulation markers.

Frequency of AE was defined as a patient’s total number of documented AEs. Severity of AE is given as the NCI-CTCAE grade. To determine a patient’s worst outcome of an AE, outcomes were ranked as follows: “Resolved”, “Ongoing/Resolving”, “Resolved with Sequelae”, “Unknown”, “Missing”, “Death”.

The analysis for this manuscript is based on an interim data-cut off on April 28, 2021.
