## Supplementary material for "High Dose Convalescent Plasma in COVID-19: Results from the Randomized Trial CAPSID": Clinical Trial Group

**The CAPSID Clinical Trial Group**

Alphabetical order

| | |
|---|---|
| Institute for Clinical Transfusion Medicine and Immunogenetics Ulm, German Red Cross Blood Transfusion Service Baden-Württemberg-Hessen and University Hospital Ulm and Institute of Transfusion Medicine, University of Ulm. | Thomas Appl, Marianne Holl, Bernd Jahrsdörfer, Sixten Körper, Ramin Lotfi, Markus Rojewski, Hubert Schrezenmeier |
| Department of Anaesthesiology and Intensive Care Medicine, University Hospital Ulm, Ulm University, Ulm, Germany | Bettina Jungwirth, Manfred Weiss |
| Department of Nephrology and Medical Intensive Care, Charité - Universitätsmedizin Berlin, corporate member of Free University Berlin, Humboldt-Universität zu Berlin, and Berlin Institute of Health, Berlin, Germany | Lucas Ernst, Jan-Matthias Kruse, Daniel Zickler, |
| Department of Anaesthesiology and Intensive Care Medicine, Phillips-University Marburg, Marburg, Germany | Tilo Koch, Thomas Wiesmann, Hinnerk Wulf |
| Clinic of Anaesthesiology, Intensive Care Medicine and Pain Therapy, University Hospital Frankfurt, Frankfurt, Germany | Elisabeth Adam, Kai Zacharowski, Sebastian Zinn |
| Institute of Virology, Charité- University Medicine Berlin, corporate member of Free University Berlin, Humboldt-Universität zu Berlin, and Berlin Institute of Health and German Centre for Infection Research, Berlin, Germany | Victor Corman, Christian Drosten, Tatjana Schwarz |
| Division of Infectious Diseases, University Hospital and Medical Center Ulm, Ulm, Germany | Beate Grüner |
| Department of Anesthesiology and Critical Care Medicine, Carl Gustav Carus University Hospital, Technische Universität Dresden, Dresden, Germany | Andreas Güldner, Peter Spieth |
| Experimental Transfusion Medicine, Technical University of Dresden, German Red Cross Blood Transfusion Service Nord-Ost gGmbH Dresden, Dresden, Germany | Torsten Tonn |
| Department of Internal Medicine V – Pneumology, Allergology, Intensive Care Medicine, Saarland University Hospital and Saarland University, Homburg, Germany | Robert Bals, Guy Danziger, Philipp M. Lepper |
| Institute for Clinical Hemostaseology and Transfusion Medicine Saarland University Hospital, Homburg, Germany | Hermann Eichler, Jan Pilch |
| Department of Internal Medicine III, Hospital of Karlsruhe, Karlsruhe, Germany | Martin Bentz, Mark Ringhoffer |
| Department of Gastroenterology, Hepatology, Pneumology and Infectious Diseases, Klinikum Stuttgart, Kriegsbergstraße 60, 70174 Stuttgart, Germany | Gregor Paul |
| Clinic of Hematology, Oncology and Palliative Care Klinikum Stuttgart, Stuttgart, Germany | Dennis Hahn |
| Clinic of Anesthesiology and Intensive Care Medicine University Medical Center of Freiburg, Germany | Johannes Kalbhenn, Julian Knörlein |
| Medical Clinic I, Klinikum Landshut, Landshut, Germany | Christian Bogner, Matthias Dollinger |
| Department of Anesthesiology and Intensive Care Medicine, University Hospital Tübingen, Tübingen, Germany | Stefanie Prohaska, Peter Rosenberger |
| Institute of Clinical and Experimental Transfusion Medicine, University Hospital Tübingen, Tübingen, Germany | Tamam Bakchoul |
| Clinic for Anesthesiology and Surgical Intensive Care Medicine, University of Mannheim, Mannheim, Germany | Thomas Kirschning, Jörg Krebs |
| Institute of Immunology and Transfusion Medicine, University Hospital Greifswald, Greifswald, Germany | Andreas Greinacher, Thomas Thiele |
| Institute of Epidemiology and Medical Biometry, Ulm University, Ulm, Germany | Benjamin Mayer |
| Institute of Transfusion Medicine and Immunology, German Red Cross Blood Transfusion Service Baden-Württemberg-Hessen, Medical Faculty of Medicine Mannheim, Heidelberg, University Mannheim, Germany | Harald Klüter, Patrick Wuchter, |
| Institute of Transfusion Medicine and Immunohematology, German Red Cross Blood Transfusion Service Baden-Württemberg – Hessen, Frankfurt, Germany | Veronika Brixner, Kay Hourfar, Michael Schmidt, Erhard Seifried |
